# Incorporating Polygenic Risk Scores and Nongenetic Risk Factors for Breast Cancer Risk Prediction among Asian Women, Results from Asia Breast Cancer Consortium

**DOI:** 10.1101/2021.09.23.21263888

**Authors:** Yaohua Yang, Ran Tao, Xiang Shu, Qiuyin Cai, Wanqing Wen, Kai Gu, Yu-Tang Gao, Ying Zheng, Sun-Seog Kweon, Min-Ho Shin, Ji-Yeob Choi, Eun-Sook Lee, Sun-Young Kong, Boyoung Park, Min Ho Park, Guochong Jia, Bingshan Li, Daehee Kang, Xiao-Ou Shu, Jirong Long, Wei Zheng

## Abstract

**Importance:** Polygenic risk scores (PRSs) have shown promises in breast cancer risk prediction; however, limited studies have been conducted among Asian women.

**Objective:** To develop breast cancer risk prediction models for Asian women incorporating PRSs and nongenetic risk factors.

**Design:** PRSs were developed using data from genome-wide association studies (GWAS) of breast cancer conducted among 123 041 Asian-ancestry women (including 18 650 cases) using three approaches (1) reported PRS for European-ancestry women; (2) breast cancer-associated single-nucleotide polymorphisms (SNPs) identified by fine-mapping of GWAS-identified risk loci; (3) genome-wide risk prediction algorithms. A nongenetic risk score (NgRS) was built including six well-established nongenetic risk factors using data from 1974 Asian women. Integrated risk scores (IRSs) were constructed using PRSs and the NgRS. PRSs were initially validated in an independent dataset including 1426 cases and 1323 controls and further evaluated, along with the NgRS and IRSs, in the second dataset including 368 cases and 736 controls nested withing a prospective cohort study.

**Setting:** Case-control and prospective cohort studies.

**Participants:** 20 444 breast cancer cases and 106 450 controls from the Asia Breast Cancer Consortium.

**Main Outcomes and Measures:** Logistic regression was used to examine associations of risk scores with breast cancer risk to estimate odds ratios (ORs) with 95% confidence intervals (CIs) and area under the receiver operating characteristic curve (AUC).

**Results:** In the prospective cohort, PRS_111,_ a PRS with 111 SNPs, developed using the fine-mapping approach showed a prediction performance comparable to a genome-wide PRS including over 855,000 SNPs. The OR per standard deviation increase of PRS_111_ was 1.67 (95% CI=1.46-1.92) with an AUC of 0.639 (95% CI=0.604-0.674). The NgRS had a limited predictive ability (AUC=0.565; 95% CI=0.529-0.601); while IRS_111_, the combination of PRS_111_ and NgRS, achieved the highest prediction accuracy (AUC=0.650; 95% CI=0.616-0.685). Compared with the average risk group (40^th^-60^th^ percentile), women in the top 5% of PRS_111_ and IRS_111_ were at a 3.84-folded (95% CI=2.30-6.46) and 4.25-folded (95% CI=2.57-7.11) elevated risk of breast cancer, respectively.

**Conclusions and Relevance:** PRSs derived using breast cancer-associated risk SNPs have similar prediction performance in Asian and European descendants. Including nongenetic risk factors in models further improved prediction accuracy. Our findings support the utility of these models in developing personalized screening and prevention strategies.

**Key Points:** *Question:* What is the performance of breast cancer risk prediction models for Asian women incorporating polygenic risk scores (PRSs) and nongenetic risk factors?

*Findings:* A 111-genetic-variant PRS developed using data of 125 790 Asian women was significantly associated with breast cancer risk in an independent case-control study nested within a prospective cohort, with an odd ratio (OR) per standard deviation increase of 1.67 (95% confidence interval [CI]=1.46-1.92) and an area under the receiver operating characteristic curve (AUC) of 0.639 (95% CI=0.604-0.674). The prediction model including this PRS and six nongenetic risk factors improved the AUC to 0.650 (95% CI=0.616-0.685).

*Meaning:* Our study provides strong supports for the utility of prediction models in identifying Asian women at high risk of breast cancer.

## Introduction

Breast cancer is the most commonly diagnosed malignancy among women worldwide.^1^ The incident rate of breast cancer has been increasing substantially in many Asian countries, although the overall rate is still significantly lower than those seen in the U.S. and many European countries.^2^ Currently, many Asian countries do not have a population-based breast cancer screening program, leading to delayed cancer diagnoses and poor survival rates.^3^ Because of the differences in breast cancer risk, screening programs currently implemented in the U.S. and European countries may not be appropriate for Asian countries. Hence, a cost-efficient, population-specific breast cancer screening strategy for Asian women is imminently needed.

In 2006, we established the Asia Breast Cancer Consortium (ABCC) to identify single nucleotide polymorphisms (SNPs) associated with breast cancer risk through genome-wide association studies (GWAS). To date, approximately 50 risk loci were identified in our studies using Asian data alone or meta-analyses of data from both Asian and European descendants.^4-12^ However, most of breast cancer risk loci were identified in GWAS conducted in European descendants.^13^ Multiple studies have attempted to aggregate effects of SNPs identified by GWAS as polygenic risk scores (PRSs) to stratify women into different breast cancer risk groups.^14-18^ The vast majority of PRSs for breast cancer were established specifically in women of European ancestry. Among them, a 313-SNP PRS showed the highest predictive ability, with an area under the receiver operating characteristic (ROC) curve (AUC) of 0.630 to 0.642.^14^ Few studies of breast cancer PRSs for Asian women were conducted and limited prediction accuracy was observed.^19- 23^ A recent study showed that the 313-SNP PRS performed better than PRSs derived from Asian data.^23^ However, in that study, the sample size was relatively small and the Asian-specific PRSs included limited number of SNPs.

In addition to genetic variations, nongenetic factors are also associated with breast cancer risk.^24^ Several studies have explored the potential of incorporating PRSs and nongenetic risk factors in improving the prediction accuracy.^18,24-27^ Among them, a recent study among European women revealed that the 313-SNP PRS was more predictive than a model including 16 nongenetic risk factors, and the best risk stratification performance was achieved when PRS and nongenetic factors were combined.^24^ However, similar studies have rarely been carried out among Asian women. In the present study, we aimed to develop and validate PRSs for Asian women using data from the largest GWAS of breast cancer ever conducted among Asian women and further evaluate the performance of risk prediction models including both PRSs and known nongenetic risk factors.

## Methods

### Study Participants

As shown in **Table 1**, the PRS development datasets included GWAS data of 20 076 breast cancer cases and 105 714 controls of Asian ancestry from the ABCC. Detailed information on the ABCC is described elsewhere.^11^ We divided these datasets to a training set (18 650 cases and 104 391 controls) for PRS derivation and a validation set (1426 cases and 1323 controls) for prediction performance evaluation (**eMethods** in the **Supplement**). For each PRS development approach, the most predictive PRS in our validation set were further evaluated in an independent case-control study nested within a prospective cohort study, comprising 368 cases and 736 individually matched controls by age (<5 years). Included in the nested case-control study were participants from the Shanghai Women’s Health Study (SWHS) and none of them had a diagnosis of any cancers at the time of enrollment (**eMethods** in the **Supplement)**.^11,28^ All studies involved in the current analyses have been approved by their respective Institutional Review Boards (IRBs). The present study has been approved by the IRB for human studies of the Vanderbilt University Medical Center.

**Table 1.** Summary of participating studies included in the current project.

|  |  |  |  |  |
| --- | --- | --- | --- | --- |
| Study | No. of cases | No. of controls | Age at enrollment <sup>a</sup> |  |
|  |  |  | Cases | Controls |
| PRS training and testing |  |  |  |  |
| *Training set* |  |  |  |  |
| SBCGS | 5384 | 6347 | 52.8 ± 9.3 | 52.1 ± 9.2 |
| HCES-Br | 274 | 273 | 49.1 ± 10.8 | 54.0 ± 7.4 |
| KPOP | 963 | 921 | - | - |
| BBJ2 | 5552 | 89 731 | - | - |
| SeBCS | 2246 | 2052 | - | - |
| BCAC-Asians | 4231 | 5067 | 54.4 ± 10.4 | 53.8 ± 10.0 |
| *Validation set* |  |  |  |  |
| SBCGS | 1426 | 1323 | 50.1 ± 11.3 | 50.6 ± 9.5 |
| *Sub-Total* | 20 076 | 105 714 |  |  |
| Prospective study |  |  |  |  |
| SWHS | 368 | 736 | 52.1 ± 8.7 | 51.6 ± 9.5 |

Abbreviations: SBCGS, Shanghai Breast Cancer Genetic Study; HCES-Br, Hwasun Cancer Epidemiology Study-Breast; KPOP, Korea Precision Oncology Program; BBJ2, The Biobank Japan Project 2; SeBCS, Seoul Breast Cancer Study; BCAC, Breast Cancer Association Consortium; SWHS, Shanghai Women's Health Study.
<sup>a</sup> Mean ± standard deviation (SD) is presented. Individual level data was not available for KPOP, BBJ2 and SeBCS.

### Genetic Data

Detailed descriptions of genetic data are provided in our recent publication and **eMethods**.^11^ Genotyping was conducted using several platforms and genotyping data imputation was performed separately by study (**eTable 1**). After quality controls and imputation, 5 947 015 SNPs were included in our analyses. GWAS was conducted within each study/sub-study and association results were combined via fixed-effects meta-analyses.

### PRS Development

We applied three approaches to develop PRSs as described below briefly and in detail in **eMethods**. PRSs were calculated using the formula: 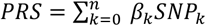, in which *SNP*_*k*_ and *β*_*k*_ represent the allelic dosage and corresponding weight of *SNP k*, and *n* is the number of SNPs used.

#### Reported PRS for European-ancestry Women

The best breast cancer PRS to date was the 313-SNP PRS among European-ancestry women.^14^ Most recently, this PRS was updated by adding 17 novel breast cancer susceptibility SNPs.^13^ Of these 330 SNPs, 263 could be found in our validation set and prospective test set, and three PRSs (PRS_263-ASN_, PRS_263-EUR_ and PRS_263-META_) were derived using weights of these SNPs from data from European-ancestry women included in the Breast Cancer Association Consortium (BCAC-European),^13^ data from Asian-ancestry women in our training set (**Table 1**), and meta-analyses of these two datasets, respectively.

#### PRSs based on SNPs selected from fine-mapping of GWAS-identified risk loci

The overall workflow of this approach is presented in **eFigure 1**. For each of the 238 independent susceptibility loci for breast cancer, ^4-12,29,30^ fine-mapping analyses were performed using summary statistics of our training set to identify SNPs that were independently associated with breast cancer risk using GCTA-COJO.^31,32^ Within each locus, a COJO-*P* threshold of 10^−5^ was used to identify independently associated risk SNPs and re-estimate weights of them on breast cancer for PRS construction. Some loci were ineligible for fine-mapping because no SNPs within them had an association with breast cancer risk at *P*<10^−5^ in our training set. Based on fine-mapping results, three PRSs were derived using (1) all SNPs selected from fine-mapping; (2) SNPs selected by fine-mapping and showing consistent association directions with *P*<.05 in the BCAC-European data; (3) SNPs in (2), plus lead SNPs from loci that were ineligible for fine-mapping but showed *P*<.05 in our training set (**eFigure 1**). We repeated the fine-mapping analyses using COJO-*P* thresholds of 10^−3^ and 10^−4^ to identify independent risk SNPs and used them to construct three sets of PRSs for each threshold following the same steps described above.

#### PRSs based on genome-wide risk prediction algorithms

LDpred, LDpred2, and PRS-CSx were used to derive genome-wide PRSs using data from the training set. The detailed description of these three algorithms can be obtained elsewhere.^33-35^ Of the 5 947 015 SNPs, indels and ambiguous SNPs were excluded by LDpred, and weights of the remaining 4 487 284 SNPs with breast cancer risk were re-evaluated. Both LDpred2 and PRS-CSx recommends using SNPs included in HapMap 3; thus, of the 5 948 258 SNPs, weights of only 855 680 HapMap 3 SNPs were re-estimated using each of these two algorithms (**eMethods** in **Supplements**).

### Models Incorporating PRSs and Nongenetic Risk Factors

Established nongenetic breast cancer risk factors included body mass index (BMI), waist-to-hip ratio (WHR), a prior diagnosis of benign breast disease, age at menarche, age at first live birth, and family history of breast cancer. An interaction term between BMI and menopause status was included in the model as BMI shows a different association with breast cancer risk by menopausal status.^19^ Data of 1974 women from the SWHS but independent from those in the prospective test set were used to estimate the weights of these six nongenetic factors and the interaction term on breast cancer risk (**eTable 2**). A logistic regression model was fitted with case/control status of breast cancer as the outcome and these eight factors as predictors. Weights estimated from this model were then used to construct a nongenetic risk score (NgRS) for each subject using the following formula: 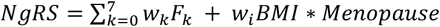, where *F*_*k*_ and *w*_*k*_ are the value and corresponding weight of factor *k*, and *w*_*i*_ is the weight of the interaction term between BMI and menopause status.

### Prediction Performance Evaluation

PRSs derived from the training set were first evaluated for their associations with breast cancer risk and prediction performance in our case-control validation set. Then, the most predictive PRSs from each PRS development approach were further appraised in our prospective test set. Finally, for the PRS showing the highest prediction accuracy in our prospective test set, and the reported European PRS,^14^ an integrated risk score (IRS) was built through incorporating each PRS with the NgRS using this formula: *IRS = PRS + NgRS*. Logistic regression was used to evaluate ORs and 95% confidence intervals (CIs) per standard deviation (SD) increase in these risk scores. Prediction performance was measured by AUCs and 95% CIs using the R function *pROC:roc*.^36^ We also investigated the utility of these scores in classifying participants with two- and three-fold increased risk compared to the average risk group (40^th^-60^th^ percentiles), through logistic regression analyses.

### Absolute Risk of Developing Breast Cancer According to PRS/IRS Percentiles

We estimated the 10-year absolute risk of developing breast cancer using the most predictive PRS in our prospective test set and the reported European PRS,^14^ and their corresponding IRSs. Considering that the prospective test set has a limited sample size (*N*=1104), which would lead to unstable OR estimates, 10 207 Chinese women (5087 cases and 5120 controls) from the whole ABCC datasets with both genetic and nongenetic data available were also included in this analysis. Logistic regression was used to estimate breast cancer ORs of different PRS/IRS percentile groups compared to the middle quintile (40%-60%) group. Then 10-year absolute risks were calculated utilizing these ORs and the incidence and mortality rates of breast cancer in Shanghai in 2017 following the strategy described previously.^19^

## Results

### Prediction Performance of PRSs

The three PRSs derived based on the reported European PRS^14^ had similar prediction performance in our case-control validation set (**Table 2; eTable 3; eTable 4**). However, in our prospective test set, PRS_263-META_ had the best prediction accuracy (AUC=0.626, 95% CI=0.592-0.661) (**Table 2**). The OR of breast cancer per SD increase of PRS_263-META_ was 1.63 (95% CI=1.43-1.87).

**Table 2.** Associations of PRSs with breast cancer risk in the validation set and prospective test set, the Asia Breast Cancer Consortium.

| PRS development methods | Validation set (1426 cases vs. 1323 controls) |  |  | Prospective test set (368 cases vs. 736 controls) |  |  |
| --- | --- | --- | --- | --- | --- | --- |
|  | OR (95% CI)<sup>a</sup> | AUC (95% CI) | *P*<sup>a</sup> | OR (95% CI)<sup>a</sup> | AUC (95% CI) | *P*<sup>a</sup> |
| **Published European PRS<sup>b</sup>** |  |  |  |  |  |  |
| PRS<sub>263</sub>-EUR | 1.42 (1.31-1.53) | 0.597 (0.575-0.618) | 2.47×10<sup>-18</sup> | 1.62 (1.42-1.85) | 0.625 (0.590-0.659) | 2.71×10<sup>-12</sup> |
| PRS<sub>263</sub>-ASN | 1.44 (1.33-1.56) | 0.601 (0.580-0.622) | 5.47×10<sup>-20</sup> | 1.58 (1.38-1.80) | 0.616 (0.582-0.651) | 1.41×10<sup>-11</sup> |
| PRS<sub>263</sub>-META | 1.44 (1.33-1.55) | 0.600 (0.579-0.621) | 1.54×10<sup>-19</sup> | 1.63 (1.43-1.87) | 0.626 (0.592-0.661) | 1.25×10<sup>-12</sup> |
| **Fine-mapping<sup>c</sup>** |  |  |  |  |  |  |
| PRS<sub>111</sub> (COJO-*P*<10<sup>-5</sup>) | 1.45 (1.34-1.57) | 0.603 (0.582-0.624) | 2.72×10<sup>-20</sup> | 1.67 (1.46-1.92) | 0.639 (0.604-0.674) | 1.28×10<sup>-13</sup> |
| PRS<sub>112</sub> (COJO-*P*<10<sup>-4</sup>) | 1.42 (1.31-1.53) | 0.597 (0.575-0.618) | 1.38×10<sup>-18</sup> | 1.63 (1.42-1.87) | 0.632 (0.597-0.667) | 1.70×10<sup>-12</sup> |
| PRS<sub>135</sub> (COJO-*P*<10<sup>-3</sup>) | 1.38 (1.28-1.49) | 0.592 (0.571-0.613) | 3.30×10<sup>-16</sup> | 1.54 (1.35-1.76) | 0.619 (0.584-0.655) | 1.55×10<sup>-10</sup> |
| **Genome-wide risk prediction algorithms<sup>d</sup>** |  |  |  |  |  |  |
| PRS<sub>LDpred</sub> (4,487,284 SNPs) | 1.44 (1.34-1.56) | 0.600 (0.579-0.621) | 4.96×10<sup>-20</sup> | 1.52 (1.34-1.74) | 0.616 (0.581-0.651) | 4.08×10<sup>-10</sup> |
| PRS<sub>LDpred2</sub> (855,680 SNPs) | 1.40 (1.29-1.51) | 0.591 (0.570-0.612) | 4.77×10<sup>-17</sup> | 1.51 (1.33-1.72) | 0.612 (0.577-0.648) | 7.47×10<sup>-10</sup> |
| PRS<sub>PRS-CSx</sub> (855,680 SNPs) | 1.51 (1.39-1.63) | 0.613 (0.592-0.634) | 3.03×10<sup>-24</sup> | 1.70 (1.49-1.95) | 0.642 (0.608-0.676) | 1.37×10<sup>-14</sup> |

PRS, polygenic risk score; OR, odds ratio; CI, confidence interval; AUC, area under the receiver operating characteristic curve.
<sup>a</sup> OR and 95% CI per SD increase in PRS and *P* values were estimated using logistic regression.
<sup>b</sup> Of the 330 SNPs included in the European-ancestry PRS reported by Zhang et al. *Nat Genet.* 2020, data on 263 SNPs were available in our validation and prospective test sets and thus included in this analysis. These PRSs were developed using weights from BCAC-European data only, Asian data only, and meta-analyses of these two datasets, respectively.
<sup>c</sup> Developed using SNPs selected from fine-mapping of Asian data and showing consistent association directions in BCAC-European data with *P*<.05. All weights were derived using Asian data.
<sup>d</sup> For each algorithm, only the most predictive PRS in the validation set is presented. Weights of SNPs from our training set were estimated using each algorithm.

To identify SNPs more specifically associated with breast cancer risk in Asian women and derive Asian-specific PRS, we performed fine-mapping analyses. At each fine-mapping threshold, three PRSs were developed (**eTable 3**) and among them, PRS_111_ showed the strongest association with breast cancer risk as well as highest prediction performance in both validation and prospective test sets (**Table 2**). This PRS was developed using 57 SNPs selected by fine-mapping and showing consistent association directions with *P*<.05 in the BCAC-European data,^13^ plus 54 lead SNPs in GWAS loci with *P*<.05 in our training set (**eFigure 1; eTable 5**). The OR for breast cancer per SD increase in PRS_111_ was 1.45 (1.34-1.57) and 1.67 (1.46-1.92), with AUCs of 0.603 (95% CI=0.582-0.624) and 0.639 (95% CI=0.604-0.674), in our case-control validation set and prospective test set, respectively (**Table 2**). Compared to the average risk group (40^th^-60^th^ percentile), women in the top 5% of PRS_111_ were at 3.84-folded (95% CI=2.30-6.46) increased risk of breast cancer. As shown in **Figures 1**. For both PRS_111_ and PRS_263-META_, distribution curves for cases were shifted to the right compared to those for controls, and the overlap was less for PRS_111_ than PRS_263-META_ (**Figures 1A** and **Figure 1C**). The difference in median percentile between cases and controls (64 vs 43) was higher for PRS_111,_ compared to PRS_263-META_ (60 vs 44) (**Figures 1B** and **Figure 1D**).

**Figure 1.**
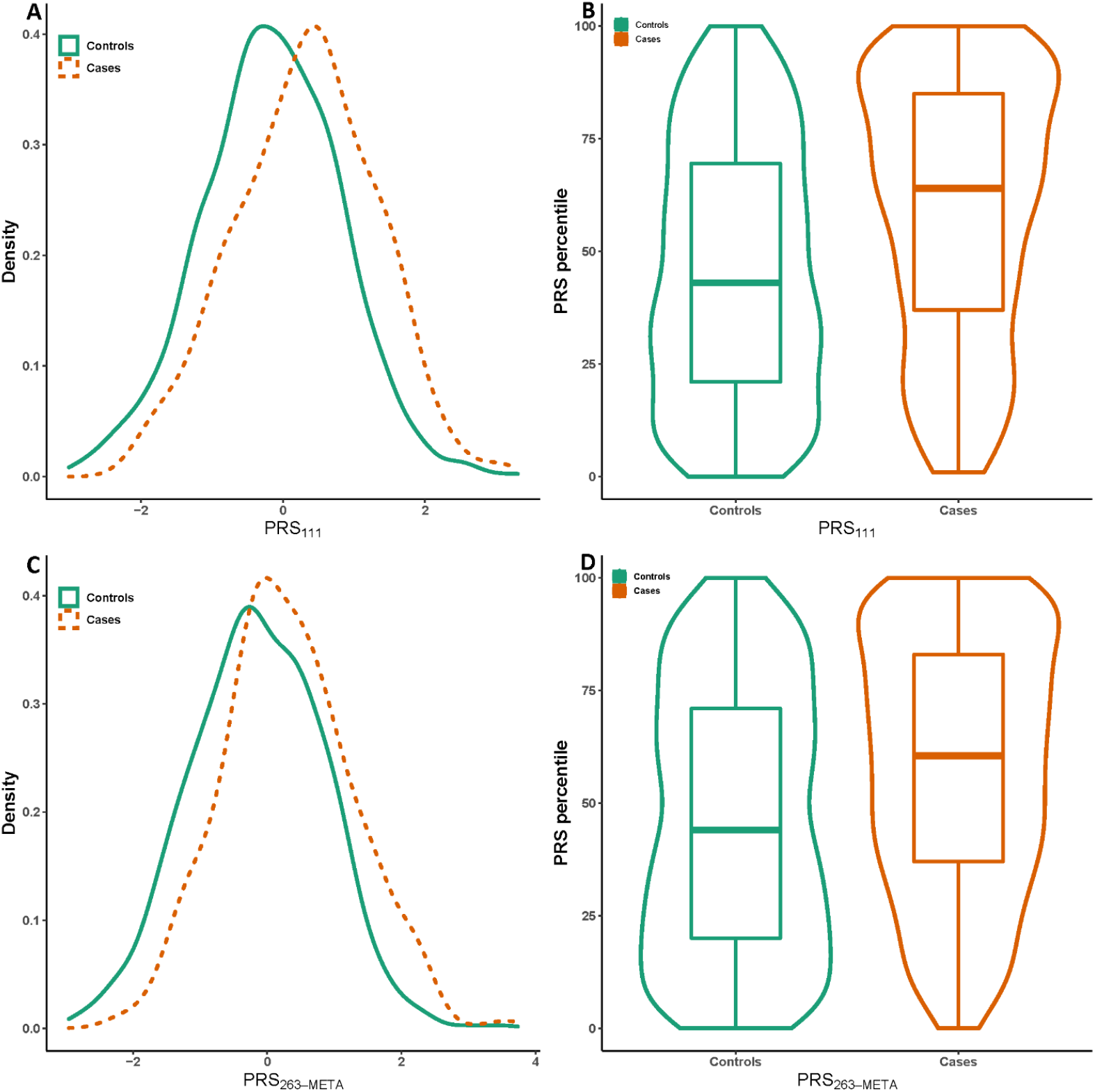
Distributions of PRS_111_ and PRS_263-META_ between breast cancer cases and controls in our prospective test set. Density plot between cases were controls for PRS_111_ (A) and PRS_263-META_ (C). Violin and boxplots between cases and controls for PRS_111_ (B) and PRS_263-META_ (D).

The most predictive PRS derived by each genome-wide risk prediction algorithm in our case-control validation set are shown in **Table 2**. Among them, PRS_PRS-CSx_ was the best-performing PRS not only in our case-control validation set, but also in our prospective test set with AUC (95% CI) of 0.613 (0.592-0.634) and 0.642 (0.608-0.676), respectively (**Table 2; eTable 3**). The OR (95% CI) for breast cancer per SD increase of this PRS in these three datasets was 1.51 (1.39-1.63) and 1.70 (1.49-1.95) respectively. Although in our prospective test set, PRS_PRS-CSx_ performed slightly better than PRS_111_ (AUC: 0.642 vs 0.639), we chose PRS_111_ as the best PRS of the present study and used it in downstream analyses because compared to PRS_PRS-CSx_, PRS_111_ used much fewer SNPs (111 vs 855 680) but had almost equal predictive ability.

### Prediction Performance of NgRS and IRSs

In our prospective test set, the NgRS was associated with breast cancer risk with an OR per SD increase of 1.29 (95% CI=1.14-1.46) with an AUC of 0.565 (95% CI=0.529-0.601) (**Table 3**). Incorporating this NgRS with PRS_111_ or PRS_263-META_, we created IRS_111_ and IRS_263-META_, respectively. In our prospective test set, IRS_111_ showed a better prediction accuracy (AUC=0.650; 95% CI=0.616-0.685; OR=1.77; 95%CI=1.55-2.04) than IRS_263-META_ (AUC=0.639; 95% CI=0.605-0.673; OR=1.72; 95% CI=1.51-1.98) **(Table 3)**. Compared to the average risk group, women in the top 5% of IRS_111_ and IRS_263-META_ were at a 4.25-folded (95% CI=2.57-7.11) and 2.79-folded (95% CI=1.70-4.63) elevated risk of breast cancer, respectively. Among all risk scores developed in the present study, IRS_111_ had the best risk stratification capability in our prospective test set. Approximately 14.0% and 38.7% of participants could be identified by IRS_111_ as having a three- and two-folded increased breast cancer risk, respectively, compared to the average risk group (**eTable 6**).

**Table 3.** Performance of risk scores in the prospective test set.

| **Risk score** | **OR (95% CI) <sup>a</sup>** | **AUC (95% CI)** | ***P* <sup>a</sup>** |
| --- | --- | --- | --- |
| NgRS <sup>b</sup> | 1.29 (1.14-1.46) | 0.565 (0.529-0.601) | 6.36×10<sup>-5</sup> |
| PRS<sub>111</sub> <sup>c</sup> | 1.67 (1.46-1.92) | 0.639 (0.604-0.674) | 1.28×10<sup>-13</sup> |
| IRS<sub>111</sub> <sup>c</sup> | 1.77 (1.55-2.04) | 0.650 (0.616-0.685) | 4.24×10<sup>-16</sup> |
| PRS<sub>263-META</sub> <sup>d</sup> | 1.63 (1.43-1.87) | 0.626 (0.592-0.661) | 1.25×10<sup>-12</sup> |
| IRS<sub>263-META</sub> <sup>d</sup> | 1.72 (1.51-1.98) | 0.639 (0.605-0.673) | 6.76×10<sup>-15</sup> |

PRS, polygenic risk score; NgRS, nongenetic risk score; IRS, integrated risk score; OR, odds ratio; CI, confidence interval; AUC, area under the receiver operating characteristic curve.
<sup>a</sup> OR and 95% CI per SD increase and *P* values was estimated using logistic regression.
<sup>b</sup> Based on weighted six nongenetic risk factors and an interaction item.
<sup>c</sup> PRS<sub>111</sub>: the best PRS derived in the present study. IRS<sub>111</sub>: the combination of PRS<sub>111</sub> and the NgRS.
<sup>d</sup> PRS<sub>263-META</sub> was derived based on meta-analysis results of Asian and BCAC-European data for 330 SNPs initially reported in European-ancestry populations (Zhang et al. *Nat Genet.* 2020). IRS<sub>263-META</sub> was the combination of PRS<sub>263-META</sub> and the NgRS.

### Absolute Risk of Developing Breast Cancer According to PRS/IRS Percentiles

Among the 10 207 Chinese women from the whole ABCC datasets, a dose-response association of breast cancer risk with percentiles of PRS_111_ or IRS_111_ was observed (**Figure 2A-B**). Compared to the average risk group, women in the top 5% of PRS_111_ and IRS_111_ were at a 3.39-folded (95% CI=2.80-4.10) and 5.22-folded (95% CI=4.37-6.25) increased risk of breast cancer, respectively; while those at the bottom 5% were at 0.30-folded (95% CI=0.23-0.39) and 0.27-folded (95% CI=0.21-0.35) decreased risk of breast cancer, respectively (**eTable 7**). The 10-year absolute risks were estimated by PRS_111_/IRS_111_ percentiles and age groups. As shown in **Figure 2C-D**, in the same percentile group, risks estimated by IRS_111_ were higher than those by PRS_111_ across age groups. For women aged 60 years, the ranges of 10-year absolute risks estimated by PRS_111_ and IRS_111_ were 0.35%-7.68% and 0.38%-14.9%, respectively. Similar results were obtained from analyses using PRS_263-META_ and IRS_263-META_ (**eFigure 2**).

**Figure 2.**
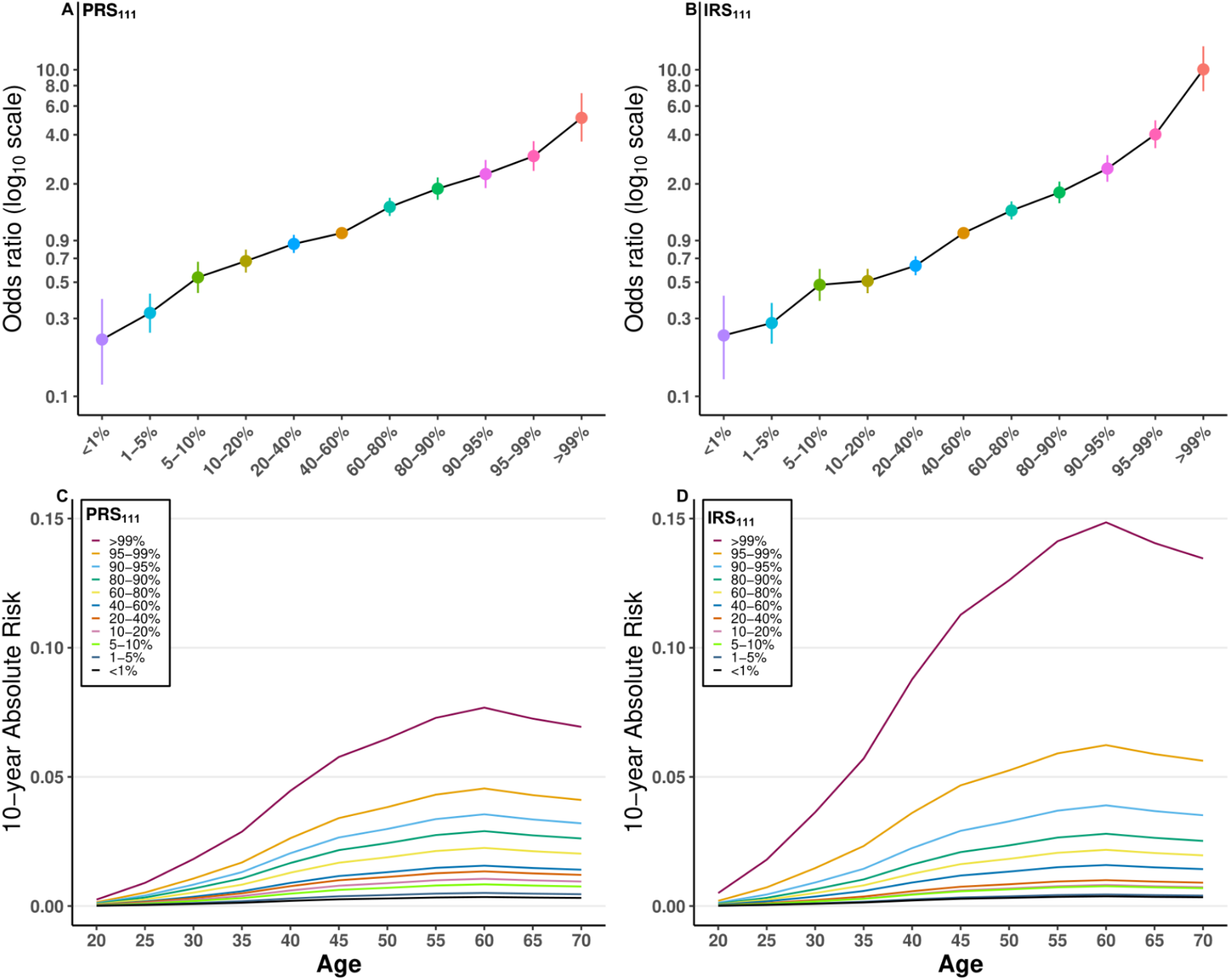
Ten-year absolute risk of developing breast cancer estimated using data from 10 207 Chinese women. ORs of breast cancer for percentiles of PRS_111_ (A) and IRS_111_ (B) compared to the average risk group (40%-60% percentiles). Ten-year absolute risk of breast cancer by percentiles of PRS_111_ (C) and IRS_111_ (D) for women in different age categories.

## Discussion

In the present study, leveraging large GWAS datasets of Asian-ancestry women, we developed PRSs for breast cancer risk using three approaches and validated their prediction performance in an independent prospective test cohort. PRS_111_, derived using the fine-mapping approach, was the best-performing PRS in this study (AUC=0.639). The prediction model incorporating PRS_111_ and six nongenetic risk factors achieved a further improved prediction accuracy (AUC=0.650). A recent study compared the predictive ability of five Asian-specific PRSs with that of the 313-SNP European PRS in a retrospective dataset of Asian women.^23^ The 313-SNP PRS was significantly more predictive (AUC=0.617) than any of the five Asian-specific PRSs (best AUC=0.586).^23^ However, because most of breast cancer risk variants were identified in GWAS conducted among European-ancestry women, the Asian-specific PRSs were derived using limited number of SNPs (n=5 to 51) in that study. In the present study, the most predictive PRSs based on these 313 SNP, PRS_263-META_, under-performed the PRS_111_, which was derived entirely using Asian data. The prediction ability of PRS_111_ in Asian women (AUC=0.639) is almost equivalent with that of the 313-SNP PRS in European-ancestry women (AUC=0.642).^14^

Most studies of prediction models incorporating PRS and nongenetic risk factors were carried out among women of European ancestry.^18,26,37-40^ In general, including nongenetic risk factors could lead to improved prediction accuracy although the magnitude of improvement is relatively small. In a recent analysis using data from a prospective cohort of Dutch women, the 313-SNP European PRS was found to have an AUC of 0.636.^39^ Incorporating this PRS with nine nongenetic risk factors improved the AUC to 0.653,^39^ similar to the level achieved in our study. In 2010, we built an Asian-specific prediction model incorporating a 12-SNP PRS, and five nongenetic risk factors, which showed an AUC of 0.629 among Chinese women.^19^ In the present study, IRS_111_, the combination of PRS_111_ and the NgRS, the aggregation of six nongenetic risk factors, outperformed both PRS_111_ and the NgRS in predicting breast cancer risk.

The strengths of this study include the use of large GWAS datasets as the training set to improve the accuracy of estimating weights of breast cancer-associated SNPs for PRS construction. Instead of using only the lead SNPs identified in original GWAS of breast cancer, we performed fine-mapping analyses to identify additional breast cancer risk SNPs specifically for Asian women. Because most of the breast-cancer associated SNPs were identified in European-ancestry populations and there are differences in genetic architectures between Asian and European-ancestry populations, we believe that this approach is necessary to conduct a PRS that is more appropriate for Asian women. In addition, state-of-the-art algorithms deriving PRSs using genome-wide SNPs were also employed in the present study and different combinations of parameters were tested for each algorithm. We demonstrated the ability of PRS-CSx in developing more predictive PRSs compared to other algorithms. Finally, the availability of both genetic and nongenetic risk factors data made it possible to establish and validate prediction models incorporating PRSs and nongenetic risk factors.

## Limitations

This study also has several limitations. First, all PRSs had better prediction performance in our prospective test set than in our case-control validation set, which may be attributed to the design of our case-control validation set, in which case and control subjects were from two different studies, which could reduce the comparability between the case and control groups. Second, the sample size of our prospective test set is relatively small, which led to relatively wide ranges of 95% CIs for ORs and AUCs. Third, we included participants from both testing and training sets to increase the sample size in the analysis of 10-year absolute risks. Although the PRSs and IRSs used for relative risk estimation were externally validated, there might still be some potential for overfitting in risk estimation. Finally, our prospective test set only includes Chinese women, hence the prediction performance of PRSs in other Asian populations could not be investigated.

In summary, using data from the largest GWAS conducted in Asian women, we demonstrated that PRSs derived using breast cancer-associated risk SNPs show similar performance in predicting breast cancer risk in Asian and European descendants. Including known nongenetic risk factors in the models could further improve the accuracy of risk prediction. Our study provides strong supports for the utility of risk prediction models in developing personalized screening and prevention strategies.

## Data Availability

Access to the ABCC data could be requested by submission of an inquiry to Dr. Wei Zheng.

## Acknowledgements

The content is solely the responsibility of the authors and does not necessarily represent the official views of the funding agents. The funders had no role in study design, data collection and analysis, decision to publish, or preparation of the manuscript. This research was primarily supported in part by the US National Institutes of Health grants R01CA235553, R01CA124558, [R01CA158473](https://grantome.com/grant/NIH/R01-CA158473-04), and R01CA148667. Sample preparation and genotyping assays at Vanderbilt were conducted at the Survey and Biospecimen Shared Resources and Vanderbilt Microarray Shared Resource, which are supported in part by the Vanderbilt-Ingram Cancer Center (P30CA068485). Data analyses were conducted using the Advanced Computing Center for Research and Education (ACCRE) at Vanderbilt University. The SeBCS was supported by the BRL (Basic Research Laboratory) program through the National Research Foundation of Korea funded by the Ministry of Education, Science and Technology (2011-0001564). KOHBRA/KOGES was supported by a grant from the National R&D Program for Cancer Control, Ministry for Health, Welfare and Family Affairs, Republic of Korea (#1020350). The KPOP was supported by Grant-in-Aid for Cancer Research and Control from the National Cancer Center of Korea (Grant Numbers 1410690 and 1710170). Studies conducted among Asian women include (Principal Investigator, grant support): the Shanghai Breast Cancer Study (W.Z. and X.-O.S., R01CA064277), the Shanghai Women’s Health Study (W.Z., R37CA070867 and UM1CA182910), the Shanghai Breast Cancer Survival Study (X.-O. S., R01CA118229), the Shanghai Endometrial Cancer Study (X.-O.S., R01CA092585, controls only), the Seoul Breast Cancer Study [D.K., BRL (Basic Research Laboratory) program through the National Research Foundation of Korea funded by the Ministry of Education, Science and Technology (2012-0000347)], the BioBank Japan Project (S.-K.L., the Ministry of Education, Culture, Sports, Sciences and Technology from the Japanese Government); the Hwasun Cancer Epidemiology Study-Breast (S.-S.K., the Biobank of Chonnam National University Hwasun Hospital, a member of the Korea Biobank Network, # 07SA2014020). The BCAC is funded by Cancer Research UK [C1287/A16563], the European Community’s Seventh Framework Programme under grant agreement 223175 [HEALTH-F2-2009-223175] (COGS).

## Notes

### Competing Interest Statement

The authors have declared no competing interest.

### Author Declarations

All studies involved in the current analyses have been approved by their respective Institutional Review Boards. The study has been approved by the internal review board for human studies of the Vanderbilt University Medical Center.

