## Supplementary files for "Incorporating Polygenic Risk Scores and Nongenetic Risk Factors for Breast Cancer Risk Prediction among Asian Women, Results from Asia Breast Cancer Consortium"

**eMethods**. Detailed Methods

**eFigure 1**. Overall workflow of the fine-mapping strategy to select SNPs for PRS development.

**eFigure 2**. Ten-year absolute risk of developing breast cancer estimated by $PRS_{263\text{-}META}$ and $IRS_{263\text{-}META}$ using data from 10 207 Chinese women.

**eTable 1**. Genotyping platforms of ABCC datasets contributing to the current study

**eTable 2**. Characteristics of participants from the prospective cohort study SWHS included in the present study

**eTable 3**. Associations of PRSs with breast cancer risk in the validation set, the prospective test set, and the combined validation and prospective test datasets

**eTable 4**. Associations of the 111 SNPs in $PRS_{111}$ with breast cancer risk in our ABCC training set and BCAC-European data

**eTable 5**. Associations with breast cancer risk for the 263 SNPs in our ABCC training set, BCAC-European data and meta-analyses

**eTable 6**. Risk stratification performance of PRSs, the NgRS, and IRSs in the prospective test set

**eTable 7**. Association with breast cancer risk for $PRS_{111}$, $IRS_{111}$, $PRS_{263\text{-}META}$ and $IRS_{263\text{-}META}$ in 10 207 Chinese women

**eReferences**

### eMethods

### Study participants

As shown in **Table 1**, the PRS development datasets included GWAS data of 20,076 breast cancer cases and 105,714 controls of Asian ancestry from the Asia Breast Cancer Consortium (ABCC), which was divided into a training set and a validation set. Detailed information on the ABCC is described elsewhere [1]. Study participants included in our training set were from six sources: the Shanghai Breast Cancer Genetics Study (SBCGS), including 11,731 Chinese women (5,384 cases and 6,347 controls) who were participants of four sub-studies; the Hwasun Cancer Epidemiology Study-Breast (HCES-Br), including 547 Korean women (274 cases and 273 controls); the Korea Precision Oncology Program (KPOP)-Breast Cancer, including 1,884 Korean women (963 cases and 921 controls); The Biobank Japan Project 2 (BBJ2), including 95,283 Japanese women (5,552 cases and 89,731 controls); the Seoul Breast Cancer Study (SeBCS), including 4,298 Korean women (2,246 cases and 2,052 controls); and eight other studies within the Breast Cancer Association Consortium (BCAC)-Asian data including 9,298 Asian-ancestry women (4,231 cases and 5,067 controls). PRSs developed using the training set were evaluated in an independent validation set from SBCGS, including 2,749 Chinese women, comprised of 1,426 cases from the Shanghai Breast Cancer Survival Study (SBCSS) [2], and 1,323 controls from the Shanghai Women's Health Study (SWHS) [3].

For each PRS development strategy, the most predictive PRSs in our validation set were further validated in an independent prospective test set comprising 368 cases and 736 individually matched controls (age ±five years old). All of these subjects were participants from the SWHS and did not have any diagnosis of any cancers at the time of enrollment [1,3]. In brief, during 1997 and 2000, the SWHS recruited approximately 75,000 adult women from urban Shanghai [3]. Incident cancer cases were identified via annual record linkage to the Shanghai Cancer Registry and in-person follow-up surveys, and confirmed by reviewing medical records [3]. All studies involved in the current analyses have been approved by their respective Institutional Review Boards and written informed consent has been obtained from all participants.

**Genotyping, imputation, quality control, and GWAS**

Detailed descriptions of genotyping, quality control (QC), and imputation procedures are described in our recent publication [1]. Genotyping was conducted using several platforms (**Table S1**) and genotyping data imputation was performed separately by study, with the 1000 Genomes Project Phase 3 (all populations) data as the reference panel. In our training set, GWAS was conducted within each study/sub-study using PLINK2.0 [4], adjusting for age, top five genetic principal components (PCs), and study (only for iCOGs and OncoArray datasets). Association results were combined via fixed-effects meta-analyses implemented in METAL [5].

In the present study, further QC steps were applied to data in our training set. First, except for the Exome BeadChip dataset that includes only ~50,000 SNPs, SNPs presented in less than half of the remaining eight datasets were excluded (**Table S1**). Then, SNPs with an imputation quality of $R^2$>0.80 in MEGA datasets and $R^2$>0.30 in all the other datasets were retained. The reasons for imposing a more stringent threshold for MEGA datasets are twofold: (1) the MEGA array contains approximately 2.10 million variants (before imputation) with an excellent genomic coverage of common variants across multi-ethnic populations; (2) data in our validation and prospective test sets were genotyped using MEGA array. For PRS construction using GWAS data from both Asian- and European-ancestry populations, we included only SNPs with a minor allele frequency (MAF) of >0.01 in both East Asian and European subjects in the 1000 Genome Project Phase 3 data. In the end, a total of 5,947,015 SNPs were included in downstream analyses.

**PRS development**

In the present study, PRSs were calculated using the formula: $PRS = \sum_{k=0}^{n} \beta_k SNP_k$, in which $SNP_k$ and $\beta_k$ represent the allelic dosage and corresponding weight of $SNP\ k$, and $n$ is the number of SNPs used.

***Reported European PRS***

For breast cancer, the best PRS to date was the one developed using 313 SNPs and their weights on breast cancer among European-ancestry women [6]. Most recently, this PRS was updated by adding 17 novel breast cancer susceptibility SNPs identified by GWAS among European-ancestry women [7]. Of these 330 SNPs, 263 could be found in our validation and prospective test sets, and three PRSs ($PRS_{263\text{-}ASN}$, $PRS_{263\text{-}EUR}$ and $PRS_{263\text{-}META}$) were derived using weights of these SNPs from our training set, BCAC-European data [7], and meta-analyses of these two datasets, respectively.

***PRSs based on SNPs selected by fine-mapping of GWAS loci***

The overall workflow of this strategy is presented in **Figure 1**. A total of 245 susceptibility loci for breast cancer have previously been identified by GWAS, including 12 identified initially in GWAS among Asian-ancestry women only,[8-14] and 28 in our recent meta-analyses conducted among Asian- and European-ancestry women [15]. Of the 245 index SNPs in these loci, seven have a linkage disequilibrium (LD) with at least one of the remaining SNPs in either East Asians or Europeans ($R^2$>0.10); hence these seven variants were excluded. For each of the remaining 238 loci, fine-mapping analysis was performed using summary statistics of our training set to identify SNPs that were independently associated with breast cancer risk via the stepwise regression strategy implemented in GCTA-COJO [16]. Genetic data of 504 subjects of East Asian ancestry included in the 1000 Genome Project Phase 3 were used as the reference for LD estimation. Within each locus, a COJO-*P* threshold of $10^{-5}$ was used to identify independent risk SNPs, weights of which on breast cancer risk were re-estimated via a joint analysis of all selected SNPs. Some loci were ineligible for fine-mapping because no SNPs within them had an association with breast cancer risk at $P<10^{-5}$ in our training set. Based on fine-mapping results, three PRSs were derived using (1) SNPs selected and weights re-estimated by fine-mapping; (2) SNPs selected by fine-mapping and showing a consistent association directions with $P<0.05$ in BCAC-European data [7], with weights re-estimated by fine-mapping; (3) SNPs and weights in (2), plus index SNPs from loci that were ineligible for fine-mapping and showed $P<0.05$ in our training set, with weights from our training set (**Figure 1**). We repeated the fine-mapping analyses using COJO-*P* thresholds of $10^{-3}$ and $10^{-4}$ to identify independent risk variants, and used them to construct three PRSs for each threshold following the same steps described above.

***PRSs based on genome-wide risk prediction algorithms***

LDpred, LDpred2, and PRS-CSx were used to derive PRSs using genome-wide SNPs. The detailed description of these three algorithms can be obtained elsewhere [17-19]. In the present study, summary statistics of associations between 5,947,015 SNPs and breast cancer were used as the input to LDpred. Genetic data of 19,257 healthy women of East Asian ancestry were used as the reference panel for pair-wise LD estimation [1]. Of the 5,947,015 SNPs, indels, ambiguous SNPs, and SNPs with MAF<0.01 were further excluded by LDpred, and weights for the remaining 4,487,284 SNPs were re-evaluated with default settings. LDpred2, a strengthened version of LDpred released recently [19], recommends using SNPs included in HapMap3 [20] data since these SNPs have sufficient coverage of the whole genome [21]. Of the 5,948,258 SNPs included in our training set, 855,680 presents in Hapmap3 data, weights of which on breast cancer risk were re-estimated using LDpred2 with default settings. Distinct from LDpred and LDpred2, PRS-CSx reevaluates the weights of genome-wide SNPs through placing a continuous shrinkage prior on them, and is capable of improve cross-population polygenic prediction through integrating summary level GWAS data and external LD reference panels from multiple populations [22]. Usage of Hapmap3 SNPs is the default setting of PRS-CSx as well, thus the input for PRS-CSx was the same as that for LDpred2, including 855,680 SNPs. Five global shrinkage parameters, 1, 0.01, $1\times10^{-4}$, $1\times10^{-6}$, and 'auto' (automatically learning from the input data), were respectively used to reevaluate weights of the 855,680 SNPs on breast cancer risk.

**Incorporation of PRSs and nongenetic risk factors**

Established nongenetic breast cancer risk factors included body mass index (BMI), menopause status, waist-to-hip ratio (WHR), a previous diagnosis of benign breast disease, age at menarche, age at first live birth, and family history of breast cancer. An interaction term between BMI and menopause status was also included [23]. Data of 1,974 women from the SWHS but independent from those in the prospective test set were used to estimate the weights of these six nongenetic factors and the interaction term on breast cancer risk. A logistic regression model was fitted with case/control status of breast cancer as the outcome and these eight factors as predictors. Weights estimated from this model were then used to construct a nongenetic risk score (NgRS) for each subject in our prospective test set using the following formula:

$$NgRS = \sum_{k=0}^{7} w_k F_k \; + \; w_i BMI * Menopause$$

In this formula, $F_k$ and $w_k$ are the value and corresponding weight of factor $k$, and $w_i$ is the weight of the interaction term between BMI and menopause status. Finally, for the PRS showing the highest prediction accuracy in our prospective test set, and the reported European PRS [7], an integrated risk score (IRS) was built through incorporating each PRS with the NgRS using this formula: $IRS = PRS + \; NgRS$.

**Prediction performance evaluation**

As mentioned above, PRSs developed using data from our training set were first evaluated in our validation set. Then, for each PRS development strategy, the most predictive PRS in our validation set was validated in our prospective test set. The NgRS and two IRSs were also validated in the prospective test set. Logistic regression was used to evaluate ORs and 95% confidence intervals (CIs) per standard deviation (SD) increase in these risk scores. Prediction performance was measured by AUCs and 95% CIs using the R function *pROC:roc* [24]. We also investigated the utility of these scores in classifying participants with two- and three-fold increased risk compared to the average risk group (40%-60% percentiles), through logistic regression analyses.

**Stratified analyses by ancestry subgroups**

Since all participants in validation and prospective test sets are Chinese, analyses stratified by ancestry subgroups could only be performed using subjects from the whole ABCC datasets. We excluded datasets that used Exome BeadChip or iCOGs for genotyping because these two platforms had relatively low genomic coverage (**Table S1**). In the remaining datasets, individual genetic data were available for 10,207 Chinese women (Affymetrix, MEGA, OncoArray datasets in SBCGS) and 2,431 Korean women (HCES-Br and KPOP) (**Table S1**). We evaluated associations with breast cancer risk and prediction performance of two PRSs, the most predictive PRS in the prospective test set and the reported European PRS [7], among these Chinese and Korean women through logistic regression and ROC analyses, respectively.

**Absolute risk of developing breast cancer according to PRS/IRS percentiles**

We estimated the 10-year absolute risk of developing breast cancer using the most predictive PRS in our prospective test set and the reported European PRS [7], and their corresponding IRSs . Considering that the prospective test set has a limited sample size ($N$=1,104), which would lead to unstable OR estimates, subjects from the whole ABCC datasets, except for the Exome BeadChip and iCOGs datasets (**Table S1**), were also included. The aforementioned 10,207 Chinese women with both genetic and nongenetic data available, including 5,087 cases and 5,120 controls, were eligible for this analysis. Logistic regression was used to estimate breast cancer ORs of different PRS/IRS percentile groups compared to the middle quintile (40%-60%) group. Then 10-year absolute risks were calculated utilizing these ORs and the incidence and mortality rates of breast cancer in Shanghai following the strategy described previously [23].

**eFigure 1**

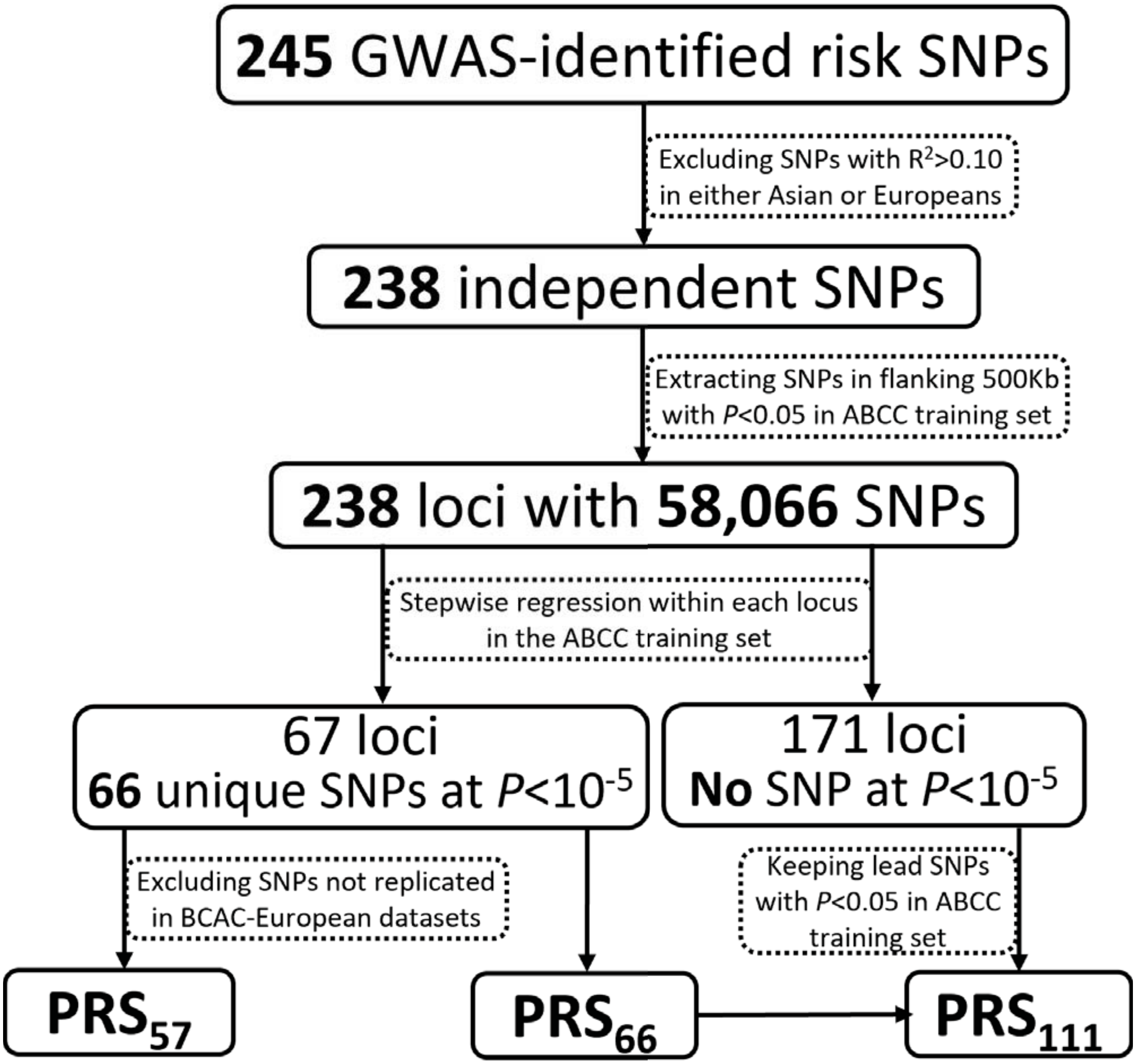

245 GWAS-identified risk SNPs
Excluding SNPs with R2>0.10 in either Asian or Europeans
238 independent SNPs
Extracting SNPs in flanking 500Kb with P<0.05 in ABCC training set
238 loci with 58,066 SNPs
Stepwise regression within each locus in the ABCC training set
67 loci 66 unique SNPs at P<10-5
171 loci No SNP at P<10-5
Excluding SNPs not replicated in BCAC-European datasets
Keeping lead SNPs with P<0.05 in ABCC training set
PRS57
PRS66
PRS111

**eFigure 2**

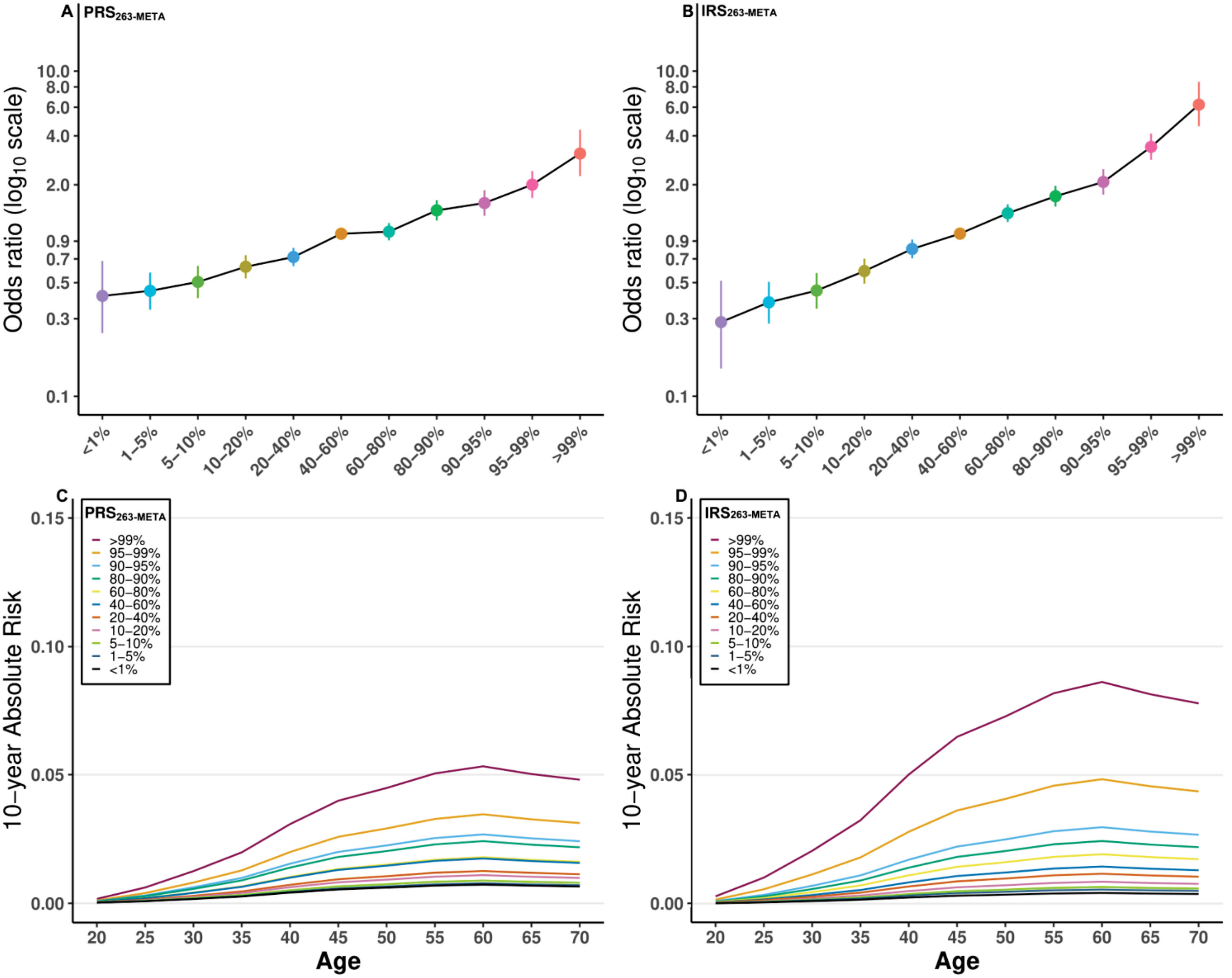

ORs of breast cancer for percentiles of $PRS_{263\text{-META}}$ (A) and $IRS_{263\text{-META}}$ (B) compared to the average risk group.

Ten-year absolute risk of breast cancer by percentiles of $PRS_{263\text{-META}}$ (C) and $IRS_{263\text{-META}}$ (D) for women in different age categories.

### eTables

#### eTable 1. Genotyping platforms of ABCC datesets contributing to the current study

| Study | Sequencing platform |
|---|---|
| **PRS development datasets** | |
| ***Training set*** | |
| SBCGS | |
| | Affymetrix GenomeWide Human SNP Array 6.0 |
| | Illumina HumanExome-12v1_A Beadchip |
| | Illumina Infinium OncoArray-500K BeadChip |
| | Illumina iSelect Genotyping Array (iCOGs) |
| HCES-Br | Illumina Multi-Ethnic Genotyping Array |
| KPOP | Illumina Multi-Ethnic Genotyping Array |
| BBJ2 | Illumina OmniExpress BeadChip |
| SeBCS | Affymetrix Genome-Wide Human SNP Array 6.0 |
| BCAC-Asian | Illumina iSelect Genotyping Array (iCOGs) |
| ***Validation set*** | |
| SBCGS | Illumina Multi-Ethnic Genotyping Array |
| **Prospective test set** | |
| SBCGS | Illumina Multi-Ethnic Genotyping Array |

Abbreviations: ABCC, Asia Breast Cancer Consortium; SBCGS, Shanghai Breast Cancer Genetic Study; HCES-Br, Hwasun Cancer Epidemiology Study-Breast; KPOP, Korea Precision Oncology Program; BBJ2, The Biobank Japan Project 2; SeBCS, Seoul Breast Cancer Study; BCAC, Breast Cancer Association Consortium

### eTable 2. Characteristics of participants from the prospective cohort study SWHS included in the present study

| Characteristics | Categories | Dataset for estimating weights of nongenetic factors - SWHS | | Prospective test set - SWHS | |
|---|---|---|---|---|---|
| | | Cases (*N*=416) | Controls (*N*=1,558) | Cases (*N*=368) | Controls (*N*=736) |
| Age at baseline interview | | | | | |
| | 40-50 | 216 (51.9%) | 815 (52.3%) | 163 (44.3%) | 365 (49.6%) |
| | 50-60 | 91 (21.9%) | 431 (27.7%) | 117 (31.8%) | 190 (25.8%) |
| | 60-70 | 109 (26.2%) | 312 (20.0%) | 88 (23.9%) | 181 (24.6%) |
| Age at menarche [a] | | 14.8 ± 1.8 | 14.9 ± 1.7 | 14.7 ± 1.6 | 14.9 ± 1.8 |
| Age at first live birth [a] | | 27.2 ± 3.9 | 26.5 ± 3.9 | 26.8 ± 4.3 | 26.2 ± 3.8 |
| Waist-to-hip ratio [a] | | 0.8 ± 0.1 | 0.8 ± 0.1 | 0.8 ± 0.1 | 0.8 ± 0.1 |
| Body mass index [a] | | 24.2 ± 3.5 | 24.0 ± 3.3 | 24.5 ± 3.4 | 24.2 ± 3.5 |
| Menopause status | | | | | |
| | Yes | 184 (44.2%) | 724 (46.5%) | 193 (52.5%) | 383 (52.0%) |
| | No | 232 (55.8%) | 834 (53.5%) | 175 (47.5%) | 353 (48.0%) |
| Family history of breast cancer | | | | | |
| | Yes | 17 (4.1%) | 29 (1.9%) | 19 (5.2%) | 17 (2.3%) |
| | No | 399 (95.9%) | 1,529 (98.1%) | 349 (94.8%) | 719 (97.7%) |
| History of benign breast disease | | | | | |
| | Yes | 114 (27.4%) | 281 (18.0%) | 63 (17.1%) | 111 (15.1%) |
| | No | 302 (72.6%) | 1,277 (82.0%) | 305 (82.9%) | 625 (84.9%) |

SWHS, Shanghai Women's Health Study.
[a] Mean ± standard deviation (SD) is presented.

**eTable 3. Associations of PRSs with breast cancer risk in the validation set, the prospective test set, and the combined validation and prospective test datasets**

| PRS development methods | Validation set (1,426 cases vs. 1,323 controls) | | | Prospective test set (368 cases vs. 736 controls) | | | Combined set (1,794 cases vs. 2,059 controls) | | |
|---|---|---|---|---|---|---|---|---|---|
| | **OR (95% CI) [a]** | **AUC (95% CI)** | ***P* [a]** | **OR (95% CI) [a]** | **AUC (95% CI)** | ***P* [a]** | **OR (95% CI) [a]** | **AUC (95% CI)** | ***P* [a]** |
| **Published European PRS [b]** | | | | | | | | | |
| $PRS_{263\text{-}EUR}$ | 1.42 (1.31-1.53) | 0.597 (0.575-0.618) | 2.47E-18 | 1.62 (1.42-1.85) | 0.625 (0.590-0.659) | 2.71E-12 | 1.48 (1.38-1.58) | 0.606 (0.588-0.623) | 1.91E-30 |
| $PRS_{263\text{-}ASN}$ | 1.44 (1.33-1.56) | 0.601 (0.580-0.622) | 5.47E-20 | 1.58 (1.38-1.80) | 0.616 (0.582-0.651) | 1.41E-11 | 1.49 (1.40-1.60) | 0.608 (0.590-0.626) | 8.53E-32 |
| $PRS_{263\text{-}META}$ | 1.44 (1.33-1.55) | 0.600 (0.579-0.621) | 1.54E-19 | 1.63 (1.43-1.87) | 0.626 (0.592-0.661) | 1.25E-12 | 1.50 (1.40-1.60) | 0.609 (0.591-0.626) | 3.81E-32 |
| **Fine-mapping [c]** | | | | | | | | | |
| **COJO-$P<10^{-3}$** | | | | | | | | | |
| $PRS_{219}$ | 1.33 (1.23-1.44) | 0.581 (0.560-0.603) | 4.07E-13 | 1.57 (1.38-1.80) | 0.619 (0.584-0.654) | 3.36E-11 | 1.39 (1.31-1.49) | 0.593 (0.575-0.611) | 4.93E-23 |
| $PRS_{120}$ | 1.36 (1.26-1.47) | 0.587 (0.566-0.608) | 4.78E-15 | 1.57 (1.38-1.80) | 0.625 (0.590-0.661) | 3.50E-11 | 1.42 (1.33-1.51) | 0.599 (0.581-0.617) | 4.84E-25 |
| $PRS_{135}$ | 1.38 (1.28-1.49) | 0.592 (0.571-0.613) | 3.30E-16 | 1.54 (1.35-1.76) | 0.619 (0.584-0.655) | 1.55E-10 | 1.42 (1.33-1.52) | 0.600 (0.582-0.618) | 9.46E-26 |
| **COJO-$P<10^{-4}$** | | | | | | | | | |
| $PRS_{99}$ | 1.33 (1.23-1.44) | 0.581 (0.560-0.602) | 2.49E-13 | 1.60 (1.40-1.83) | 0.622 (0.587-0.657) | 4.77E-12 | 1.40 (1.31-1.50) | 0.593 (0.575-0.611) | 1.53E-23 |
| $PRS_{73}$ | 1.37 (1.27-1.48) | 0.588 (0.567-0.609) | 1.85E-15 | 1.60 (1.40-1.83) | 0.630 (0.595-0.665) | 6.03E-12 | 1.43 (1.34-1.53) | 0.600 (0.582-0.618) | 5.29E-26 |
| $PRS_{112}$ | 1.42 (1.31-1.53) | 0.597 (0.575-0.618) | 1.38E-18 | 1.63 (1.42-1.87) | 0.632 (0.597-0.667) | 1.70E-12 | 1.47 (1.38-1.57) | 0.607 (0.589-0.624) | 4.97E-30 |
| **COJO-$P<10^{-5}$** | | | | | | | | | |
| $PRS_{66}$ | 1.37 (1.27-1.48) | 0.590 (0.569-0.611) | 1.30E-15 | 1.66 (1.45-1.90) | 0.634 (0.600-0.669) | 1.91E-13 | 1.45 (1.35-1.55) | 0.603 (0.585-0.621) | 9.91E-28 |
| $PRS_{57}$ | 1.38 (1.28-1.49) | 0.591 (0.570-0.612) | 4.87E-16 | 1.66 (1.45-1.91) | 0.641 (0.606-0.675) | 2.16E-13 | 1.45 (1.36-1.55) | 0.605 (0.587-0.623) | 4.12E-28 |
| $PRS_{111}$ | 1.45 (1.34-1.57) | 0.603 (0.582-0.624) | 2.72E-20 | 1.67 (1.46-1.92) | 0.639 (0.604-0.674) | 1.28E-13 | 1.51 (1.41-1.61) | 0.614 (0.596-0.631) | 4.22E-33 |
| **Genome-wide Bayesian algorithms [d]** | | | | | | | | | |
| $PRS_{LDpred}$ (4,487,284 SNPs) | 1.44 (1.34-1.56) | 0.600 (0.579-0.621) | 4.96E-20 | 1.52 (1.34-1.74) | 0.616 (0.581-0.651) | 4.08E-10 | 1.47 (1.37-1.57) | 0.604 (0.586-0.622) | 2.90E-29 |
| $PRS_{LDpred2}$ (855,680 SNPs) | 1.40 (1.29-1.51) | 0.591 (0.570-0.612) | 4.77E-17 | 1.51 (1.33-1.72) | 0.612 (0.577-0.648) | 7.47E-10 | 1.43 (1.34-1.52) | 0.597 (0.579-0.615) | 7.17E-26 |
| $PRS_{PRS\text{-}CSx}$ (855,680 SNPs) | 1.51 (1.39-1.63) | 0.613 (0.592-0.634) | 3.03E-24 | 1.70 (1.49-1.95) | 0.642 (0.608-0.676) | 1.37E-14 | 1.55 (1.45-1.66) | 0.620 (0.602-0.637) | 2.08E-37 |

PRS, polygenic risk score; OR, odds ratio; CI, confidence interval; AUC, aera under the receiver operating characteristic curve.

[a] OR and 95% CI per standard deviation (SD) increase in PRS and *P* values was estimated using logistic regression.

[b] The 330-SNP European PRS reported by Zhang et al. *Nat Genet*. 2020. Based on 263 of the 330 SNPs that were available in our validation and prospective test sets, three PRSs were developed using weights from our training set, BCAC-European data, and meta-analyses these two datasets, respectively.

[c] At each COJO-*P* threshold, three PRSs were developed, respectively using (1) SNPs selected and weights re-estimated by fine-mapping; (2) SNPs from (1) and showing consistent association directions in BCAC-European data with $P<0.05$, with weights from fine-mapping; (3) SNPs and weights in (2), adding index SNPs in loci that were ineligible for fine-mapping and showing $P<0.05$ in our training set, and weights from our training set.

[d] For each algorithm, only the most predictive PRS in the validation set is presented. Weights of SNPs from our training set were re-estimated by each algorithm.

**eTable 4. Associations with breast cancer risk for the 263 SNPs in our ABCC training set, BCAC-European data and meta-analyses**

| RSID | Chr | Position (GRCh37) | Effect allele | Non-effect allele | ABCC training set | | | BCAC-European data | | | Meta-analyses | |
|---|---|---|---|---|---|---|---|---|---|---|---|---|
| | | | | | Effect allele frequency (%) | Effect size | *P* | Effect allele frequency (%) | Effect size [b] | *P* [b] | Effect size | *P* |
| rs707475 | 1 | 7,917,076 | A | G | 19.35 | 0.00 | 1.00 | 41.95 | -0.03 | 1.55E-07 | -0.03 | 1.01E-06 |
| rs616488 | 1 | 10,566,215 | G | A | 31.75 | -0.05 | 2.52E-04 | 32.60 | -0.06 | 9.62E-21 | -0.06 | 1.70E-23 |
| rs2992756 | 1 | 18,807,339 | C | T | 85.71 | -0.04 | 6.83E-03 | 48.81 | -0.05 | 2.67E-17 | -0.05 | 7.10E-19 |
| rs4233486 | 1 | 41,380,440 | T | C | 64.88 | 0.01 | 0.49 | 62.82 | 0.04 | 3.81E-08 | 0.03 | 1.57E-07 |
| rs17426269 | 1 | 88,156,923 | A | G | 0.10 | 0.10 | 0.41 | 12.43 | 0.04 | 3.87E-07 | 0.04 | 3.04E-07 |
| rs2151842 | 1 | 88,428,199 | A | C | 12.40 | -0.02 | 0.18 | 26.94 | -0.04 | 4.88E-09 | -0.04 | 2.80E-09 |
| rs612683 | 1 | 100,880,328 | T | A | 51.98 | -0.01 | 0.45 | 38.17 | 0.03 | 3.62E-07 | 0.02 | 2.43E-05 |
| rs7513707 | 1 | 114,445,880 | A | G | 58.63 | 0.02 | 0.19 | 19.18 | 0.06 | 3.54E-12 | 0.04 | 4.94E-11 |
| rs12406858 | 1 | 118,141,492 | C | A | 46.03 | 0.02 | 0.11 | 27.24 | 0.04 | 2.14E-07 | 0.03 | 1.10E-07 |
| rs637868 | 1 | 120,257,110 | C | T | 89.78 | -0.04 | 0.11 | 53.58 | 0.04 | 5.89E-09 | 0.03 | 2.10E-07 |
| rs11249433 | 1 | 121,280,613 | G | A | 2.98 | 0.09 | 0.01 | 43.14 | 0.10 | 1.21E-57 | 0.10 | 6.15E-59 |
| rs143384623 | 1 | 145,604,302 | CT | C | 16.57 | -0.01 | 0.74 | 34.00 | -0.04 | 3.16E-10 | -0.04 | 1.28E-09 |
| rs11205303 | 1 | 149,906,413 | C | T | 28.37 | 0.03 | 0.06 | 36.88 | 0.05 | 5.07E-16 | 0.05 | 2.55E-16 |
| rs12091730 | 1 | 155,556,971 | A | G | 66.67 | 0.05 | 1.71E-03 | 22.86 | 0.05 | 1.83E-12 | 0.05 | 1.27E-14 |
| rs11463354 | 1 | 172,328,767 | TA | T | 13.10 | 0.01 | 0.53 | 32.31 | -0.03 | 4.30E-05 | -0.02 | 1.84E-04 |
| rs6686987 | 1 | 202,184,600 | T | C | 25.89 | -0.03 | 0.06 | 42.64 | -0.01 | 0.04 | -0.01 | 9.45E-03 |
| rs7514172 | 1 | 203,770,448 | A | T | 32.44 | 0.06 | 8.86E-06 | 24.45 | 0.05 | 1.78E-12 | 0.05 | 9.19E-17 |
| rs2785646 | 1 | 208,076,291 | A | G | 2.18 | -0.01 | 0.84 | 36.18 | -0.03 | 5.12E-08 | -0.03 | 5.56E-08 |
| rs11117758 | 1 | 217,220,574 | A | G | 3.87 | -0.03 | 0.32 | 22.76 | -0.04 | 4.28E-09 | -0.04 | 2.98E-09 |
| rs11118563 | 1 | 220,671,050 | T | C | 35.22 | -0.01 | 0.67 | 22.86 | 0.03 | 6.13E-05 | 0.02 | 1.06E-03 |
| rs72755295 | 1 | 242,034,263 | G | A | 0.00 | 0.45 | 0.27 | 3.78 | 0.12 | 1.38E-12 | 0.12 | 1.03E-12 |
| rs6743383 | 2 | 19,315,675 | A | T | 59.03 | -0.04 | 1.04E-03 | 59.84 | -0.04 | 1.91E-13 | -0.04 | 8.50E-16 |
| rs6725517 | 2 | 25,129,473 | G | A | 42.16 | -0.02 | 0.15 | 39.36 | -0.04 | 1.06E-09 | -0.03 | 1.29E-09 |
| rs12472404 | 2 | 29,179,452 | C | G | 80.06 | 0.00 | 0.84 | 22.76 | 0.00 | 0.77 | 0.00 | 0.73 |
| rs9712235 | 2 | 67,881,757 | A | G | 81.05 | 0.00 | 0.83 | 75.94 | -0.04 | 4.78E-08 | -0.03 | 4.46E-07 |
| rs4602255 | 2 | 69,392,128 | A | G | 89.29 | 0.02 | 0.44 | 47.71 | 0.04 | 1.95E-09 | 0.03 | 2.01E-09 |
| rs6756513 | 2 | 70,172,587 | A | G | 29.46 | -0.05 | 7.37E-05 | 27.34 | -0.04 | 1.03E-07 | -0.04 | 5.30E-11 |
| rs1036759 | 2 | 88,358,825 | C | G | 27.08 | 0.02 | 0.32 | 32.41 | 0.03 | 1.64E-04 | 0.02 | 1.20E-04 |
| rs6746250 | 2 | 121,058,254 | G | A | 40.28 | -0.02 | 0.22 | 70.78 | -0.03 | 1.38E-07 | -0.03 | 1.64E-07 |
| rs17625845 | 2 | 121,089,731 | C | T | 7.04 | 0.01 | 0.61 | 22.07 | -0.04 | 1.89E-07 | -0.03 | 2.42E-06 |
| rs10164550 | 2 | 121,159,205 | A | G | 12.90 | -0.01 | 0.51 | 37.38 | -0.05 | 9.87E-14 | -0.04 | 3.57E-13 |

| | | | | | | | | | | | | |
|---|---|---|---|---|---|---|---|---|---|---|---|---|
| rs10179592 | 2 | 121,246,568 | C | T | 79.07 | 0.08 | 7.55E-08 | 89.76 | 0.10 | 7.23E-23 | 0.09 | 4.79E-29 |
| rs17726078 | 2 | 172,974,566 | G | C | 18.55 | -0.01 | 0.37 | 45.63 | -0.04 | 1.51E-11 | -0.04 | 3.45E-11 |
| rs1550622 | 2 | 174,212,910 | G | A | 99.50 | 0.14 | 0.07 | 84.69 | 0.05 | 6.75E-10 | 0.05 | 2.46E-10 |
| rs2356656 | 2 | 192,381,934 | T | C | 94.64 | -0.03 | 0.17 | 85.39 | 0.02 | 0.05 | 0.01 | 0.17 |
| rs10197246 | 2 | 202,204,741 | C | T | 70.63 | -0.09 | 4.38E-13 | 70.97 | -0.06 | 4.38E-17 | -0.06 | 3.79E-27 |
| rs4442975 | 2 | 217,920,769 | T | G | 89.88 | -0.08 | 3.30E-05 | 49.11 | -0.13 | 1.03E-109 | -0.13 | 8.91E-112 |
| rs11693806 | 2 | 218,292,158 | G | C | 38.19 | -0.06 | 1.27E-06 | 71.47 | -0.07 | 2.75E-27 | -0.07 | 3.39E-32 |
| rs3791977 | 2 | 218,714,845 | A | G | 27.88 | -0.02 | 0.10 | 34.99 | -0.03 | 4.76E-07 | -0.03 | 1.41E-07 |
| rs6762558 | 3 | 4,742,251 | G | A | 6.55 | 0.05 | 0.02 | 35.19 | 0.05 | 3.73E-19 | 0.05 | 2.16E-20 |
| rs1375631 | 3 | 16,778,867 | G | A | 2.08 | 0.05 | 0.32 | 49.80 | 0.03 | 6.79E-09 | 0.03 | 4.19E-09 |
| rs552647 | 3 | 27,353,716 | A | C | 26.59 | 0.07 | 1.65E-07 | 53.28 | 0.10 | 1.86E-67 | 0.10 | 3.25E-72 |
| rs62255657 | 3 | 27,388,664 | G | C | 13.29 | 0.07 | 9.79E-06 | 29.42 | 0.10 | 5.01E-48 | 0.09 | 6.14E-52 |
| rs17838698 | 3 | 30,684,907 | T | C | 69.15 | 0.04 | 2.39E-03 | 28.93 | 0.05 | 4.40E-14 | 0.05 | 5.74E-16 |
| rs56387622 | 3 | 46,888,198 | C | T | 14.68 | -0.05 | 3.34E-03 | 10.04 | -0.09 | 9.68E-20 | -0.08 | 1.76E-20 |
| rs371314787 | 3 | 49,709,912 | CT | C | 6.35 | -0.02 | 0.55 | 30.91 | -0.02 | 6.02E-04 | -0.02 | 5.04E-04 |
| rs2886671 | 3 | 59,373,745 | T | C | 67.76 | -0.01 | 0.66 | 40.46 | -0.04 | 4.25E-08 | -0.03 | 2.56E-07 |
| rs147250346 | 3 | 63,887,449 | TTG | T | 13.99 | 0.02 | 0.42 | 14.31 | 0.06 | 1.60E-12 | 0.06 | 7.90E-12 |
| rs9825432 | 3 | 71,620,370 | G | T | 3.27 | -0.02 | 0.61 | 67.79 | -0.04 | 4.88E-09 | -0.04 | 5.00E-09 |
| rs639355 | 3 | 99,403,877 | A | G | 47.92 | 0.00 | 0.80 | 51.89 | -0.03 | 1.12E-08 | -0.03 | 1.66E-07 |
| rs376397524 | 3 | 141,112,859 | C | CTT | 7.44 | 0.05 | 0.07 | 40.06 | 0.05 | 4.38E-16 | 0.05 | 8.54E-17 |
| rs58058861 | 3 | 172,285,237 | A | G | 32.74 | 0.03 | 0.03 | 17.89 | 0.05 | 2.40E-10 | 0.04 | 2.95E-11 |
| rs9882792 | 3 | 189,774,456 | T | C | 10.42 | 0.01 | 0.63 | 20.87 | -0.03 | 4.48E-05 | -0.03 | 2.03E-04 |
| rs495367 | 4 | 1,986,972 | G | A | 28.87 | -0.01 | 0.54 | 30.62 | 0.04 | 5.28E-07 | 0.03 | 1.91E-05 |
| rs10012017 | 4 | 38,784,633 | T | G | 48.12 | 0.02 | 0.21 | 27.83 | 0.05 | 7.66E-13 | 0.04 | 8.14E-12 |
| rs532161833 | 4 | 84,370,124 | TA | TAA | 69.35 | -0.03 | 0.08 | 51.79 | -0.04 | 2.50E-12 | -0.04 | 7.52E-13 |
| rs17014016 | 4 | 89,240,476 | A | G | 1.19 | 0.09 | 0.13 | 42.84 | 0.04 | 2.47E-09 | 0.04 | 1.19E-09 |
| rs62331150 | 4 | 106,069,013 | T | G | 63.59 | 0.00 | 0.71 | 21.17 | 0.04 | 4.09E-10 | 0.03 | 2.37E-08 |
| rs147399132 | 4 | 126,752,992 | AAT | A | 35.91 | 0.01 | 0.46 | 50.60 | -0.03 | 5.23E-07 | -0.03 | 8.14E-06 |
| rs56039025 | 4 | 143,467,195 | T | C | 2.68 | 0.00 | 0.93 | 12.03 | -0.04 | 1.93E-05 | -0.04 | 2.88E-05 |
| rs138786872 | 4 | 151,218,296 | C | CATATTT | 26.09 | -0.01 | 0.52 | 62.13 | 0.03 | 1.98E-07 | 0.03 | 7.69E-06 |
| rs28436676 | 4 | 175,842,495 | A | G | 23.81 | -0.07 | 1.19E-07 | 10.83 | -0.10 | 5.54E-27 | -0.09 | 1.43E-32 |
| rs62334414 | 4 | 175,847,436 | A | C | 1.79 | 0.04 | 0.31 | 35.09 | 0.05 | 3.02E-15 | 0.05 | 1.90E-15 |
| rs10069690 | 5 | 1,279,790 | T | C | 16.87 | 0.06 | 3.52E-03 | 27.63 | 0.06 | 1.76E-18 | 0.06 | 2.36E-20 |
| rs3215401 | 5 | 1,296,255 | AG | A | 38.39 | -0.03 | 0.02 | 28.83 | -0.07 | 1.13E-23 | -0.06 | 5.12E-24 |
| rs4866496 | 5 | 2,777,029 | A | G | 76.79 | 0.03 | 0.02 | 43.04 | 0.03 | 4.22E-07 | 0.03 | 2.19E-08 |
| rs17611291 | 5 | 16,231,194 | C | G | 15.77 | 0.02 | 0.32 | 56.06 | -0.04 | 2.89E-12 | -0.04 | 5.26E-10 |
| rs4613718 | 5 | 44,649,944 | T | C | 55.26 | 0.11 | 3.90E-19 | 61.63 | 0.05 | 5.85E-16 | 0.06 | 6.41E-29 |
| rs10941679 | 5 | 44,706,498 | G | A | 48.71 | 0.10 | 1.96E-17 | 23.26 | 0.13 | 3.82E-84 | 0.13 | 7.98E-99 |
| rs17343002 | 5 | 44,853,593 | C | G | 6.15 | -0.12 | 3.12E-07 | 31.11 | -0.05 | 7.12E-14 | -0.05 | 9.53E-18 |
| rs199562199 | 5 | 52,679,539 | CA | C | 19.54 | 0.02 | 0.28 | 11.23 | 0.05 | 1.81E-07 | 0.05 | 2.54E-07 |
| rs553874618 | 5 | 55,662,540 | CT | C | 36.81 | -0.02 | 0.09 | 39.76 | -0.03 | 3.61E-06 | -0.03 | 8.94E-07 |

| | | | | | | | | | | | | |
|---|---|---|---|---|---|---|---|---|---|---|---|---|
| rs889310 | 5 | 55,965,167 | T | C | 60.81 | 0.04 | 7.96E-04 | 58.15 | 0.04 | 3.32E-10 | 0.04 | 1.11E-12 |
| rs16886165 | 5 | 56,023,083 | G | T | 34.03 | 0.06 | 7.29E-07 | 15.01 | 0.17 | 3.28E-104 | 0.14 | 2.26E-97 |
| rs76250845 | 5 | 56,042,972 | T | C | 11.11 | 0.18 | 1.33E-18 | 4.87 | 0.20 | 3.48E-55 | 0.20 | 6.80E-72 |
| rs11949391 | 5 | 56,045,081 | C | T | 5.16 | -0.06 | 0.07 | 14.02 | -0.09 | 1.50E-27 | -0.09 | 4.89E-28 |
| rs113778879 | 5 | 58,241,712 | T | C | 65.28 | 0.00 | 0.94 | 57.36 | -0.04 | 1.70E-10 | -0.03 | 6.33E-09 |
| rs138044103 | 5 | 67,424,121 | CTG | C | 78.97 | 0.03 | 0.07 | 48.01 | 0.02 | 1.16E-04 | 0.02 | 2.13E-05 |
| rs3010266 | 5 | 71,965,007 | A | G | 11.41 | -0.05 | 9.64E-03 | 23.16 | -0.04 | 7.44E-07 | -0.04 | 2.85E-08 |
| rs157557 | 5 | 73,234,583 | C | T | 42.16 | 0.01 | 0.48 | 32.01 | -0.03 | 5.16E-05 | -0.02 | 1.07E-03 |
| rs144028731 | 5 | 77,155,397 | G | GT | 17.06 | -0.03 | 0.12 | 37.67 | -0.03 | 5.21E-06 | -0.03 | 1.49E-06 |
| rs34525310 | 5 | 79,180,995 | GA | G | 38.69 | -0.01 | 0.39 | 15.01 | 0.03 | 3.55E-04 | 0.02 | 0.01 |
| rs146817970 | 5 | 81,512,947 | T | TA | 0.00 | -0.23 | 0.17 | 23.16 | -0.05 | 3.91E-14 | -0.05 | 2.64E-14 |
| rs332529 | 5 | 90,789,470 | A | G | 48.51 | -0.07 | 8.33E-08 | 14.91 | -0.06 | 6.63E-14 | -0.06 | 3.17E-20 |
| rs17157372 | 5 | 104,300,273 | T | G | 5.95 | 0.00 | 0.88 | 18.49 | -0.02 | 2.90E-03 | -0.02 | 4.25E-03 |
| rs335160 | 5 | 122,478,676 | A | C | 61.90 | 0.00 | 1.00 | 77.34 | -0.03 | 3.22E-06 | -0.02 | 4.64E-05 |
| rs1428387 | 5 | 122,705,244 | T | C | 8.43 | 0.03 | 0.14 | 2.68 | 0.09 | 3.91E-07 | 0.06 | 1.56E-06 |
| rs6860806 | 5 | 131,640,536 | G | A | 30.46 | 0.01 | 0.46 | 52.88 | 0.03 | 2.66E-08 | 0.03 | 7.62E-08 |
| rs6596100 | 5 | 132,407,058 | T | C | 9.13 | -0.01 | 0.56 | 23.16 | -0.04 | 1.70E-08 | -0.04 | 3.98E-08 |
| rs1432679 | 5 | 158,244,083 | T | C | 38.89 | -0.07 | 9.14E-08 | 55.27 | -0.07 | 1.56E-30 | -0.07 | 9.11E-37 |
| rs10074269 | 5 | 169,591,460 | C | T | 44.35 | 0.02 | 0.11 | 34.10 | 0.04 | 1.09E-09 | 0.04 | 8.05E-10 |
| rs6864691 | 5 | 173,358,154 | A | G | 29.17 | -0.01 | 0.68 | 40.16 | 0.03 | 3.29E-06 | 0.02 | 4.87E-05 |
| rs418053 | 6 | 13,713,366 | C | G | 38.89 | -0.04 | 6.77E-04 | 58.35 | -0.05 | 1.56E-15 | -0.05 | 5.21E-18 |
| rs543824204 | 6 | 20,537,845 | C | CA | 31.65 | -0.02 | 0.11 | 46.92 | -0.04 | 1.61E-09 | -0.04 | 6.29E-10 |
| rs9358466 | 6 | 21,923,810 | C | T | 28.87 | -0.02 | 0.13 | 45.23 | -0.04 | 4.04E-09 | -0.03 | 2.18E-09 |
| rs17215231 | 6 | 33,239,869 | T | C | 2.38 | -0.01 | 0.74 | 7.46 | -0.03 | 0.01 | -0.03 | 0.01 |
| rs111342015 | 6 | 43,227,141 | A | G | 0.40 | 0.13 | 0.06 | 6.56 | -0.05 | 1.27E-07 | -0.05 | 7.59E-07 |
| rs574103382 | 6 | 82,263,549 | A | AAT | 42.26 | 0.02 | 0.20 | 42.84 | 0.05 | 1.73E-14 | 0.04 | 9.48E-14 |
| rs73754909 | 6 | 87,803,819 | C | T | 25.50 | 0.00 | 0.77 | 29.03 | 0.03 | 5.93E-05 | 0.02 | 4.53E-04 |
| rs55941023 | 6 | 130,341,728 | CT | C | 93.15 | 0.02 | 0.51 | 70.78 | 0.04 | 3.92E-11 | 0.04 | 6.28E-11 |
| rs2121348 | 6 | 149,595,505 | C | T | 42.96 | -0.08 | 2.64E-12 | 18.39 | -0.04 | 3.66E-08 | -0.05 | 4.51E-17 |
| rs6913578 | 6 | 151,949,806 | C | A | 33.93 | 0.20 | 1.51E-55 | 29.03 | 0.09 | 6.28E-47 | 0.11 | 3.17E-88 |
| rs60954078 | 6 | 151,955,914 | G | A | 31.25 | 0.20 | 4.33E-50 | 7.95 | 0.18 | 8.74E-56 | 0.19 | 8.57E-104 |
| rs851984 | 6 | 152,023,191 | A | G | 9.82 | 0.11 | 1.23E-09 | 41.75 | 0.06 | 3.31E-21 | 0.06 | 9.60E-28 |
| rs6904031 | 6 | 152,055,978 | T | A | 6.85 | 0.14 | 7.16E-11 | 5.96 | 0.14 | 4.38E-29 | 0.14 | 2.42E-38 |
| rs910416 | 6 | 152,432,902 | T | C | 57.34 | 0.04 | 1.04E-03 | 54.77 | 0.06 | 4.48E-27 | 0.06 | 1.21E-28 |
| rs9364472 | 6 | 169,006,947 | G | C | 36.90 | 0.01 | 0.55 | 54.87 | -0.02 | 1.54E-04 | -0.02 | 1.23E-03 |
| rs6940159 | 6 | 170,332,621 | C | T | 14.78 | 0.07 | 2.43E-05 | 59.44 | 0.03 | 1.10E-07 | 0.04 | 9.23E-11 |
| rs7971 | 7 | 21,940,960 | G | A | 16.77 | -0.01 | 0.72 | 35.69 | -0.04 | 1.69E-08 | -0.03 | 8.81E-08 |
| rs289997 | 7 | 25,569,548 | T | C | 24.70 | 0.01 | 0.67 | 15.81 | -0.04 | 5.54E-07 | -0.03 | 2.17E-05 |
| rs13244925 | 7 | 55,192,256 | C | A | 68.85 | -0.03 | 0.04 | 53.38 | -0.03 | 2.41E-06 | -0.03 | 2.58E-07 |
| rs17268829 | 7 | 94,113,799 | C | T | 28.47 | 0.05 | 5.55E-04 | 28.83 | 0.05 | 2.58E-13 | 0.05 | 6.12E-16 |
| rs111963714 | 7 | 99,948,655 | G | T | 3.08 | 0.00 | 0.94 | 19.98 | 0.03 | 1.17E-05 | 0.03 | 1.84E-05 |

| | | | | | | | | | | | | |
|---|---|---|---|---|---|---|---|---|---|---|---|---|
| rs71559437 | 7 | 101,552,440 | A | G | 7.04 | -0.05 | 0.13 | 11.03 | -0.06 | 1.19E-09 | -0.06 | 3.80E-10 |
| rs7800548 | 7 | 102,481,842 | C | T | 57.64 | 0.00 | 0.71 | 31.81 | 0.03 | 1.44E-07 | 0.03 | 6.38E-06 |
| rs12706954 | 7 | 130,656,911 | T | C | 24.31 | -0.01 | 0.49 | 35.39 | -0.04 | 1.44E-11 | -0.04 | 7.99E-11 |
| rs68056147 | 7 | 130,674,481 | A | G | 26.39 | 0.05 | 9.85E-04 | 29.72 | 0.05 | 2.97E-15 | 0.05 | 1.24E-17 |
| rs5887960 | 7 | 139,943,702 | C | CT | 54.17 | 0.05 | 8.48E-05 | 53.68 | 0.06 | 5.64E-18 | 0.05 | 2.38E-21 |
| rs66823261 | 8 | 170,692 | C | T | 20.44 | 0.02 | 0.20 | 20.38 | 0.04 | 2.37E-06 | 0.03 | 1.41E-06 |
| rs1028016 | 8 | 23,447,496 | G | A | 84.52 | 0.02 | 0.32 | 67.30 | -0.03 | 1.84E-05 | -0.02 | 1.75E-04 |
| rs310295 | 8 | 23,663,653 | A | C | 29.96 | 0.03 | 0.02 | 39.56 | 0.03 | 1.43E-06 | 0.03 | 9.69E-08 |
| rs13256025 | 8 | 25,831,778 | T | C | 1.39 | 0.02 | 0.62 | 22.56 | 0.04 | 1.41E-08 | 0.04 | 1.53E-08 |
| rs9693444 | 8 | 29,509,616 | C | A | 70.04 | -0.06 | 2.20E-06 | 65.41 | -0.06 | 6.82E-21 | -0.06 | 8.31E-26 |
| rs13365225 | 8 | 36,858,483 | G | A | 31.25 | -0.06 | 2.65E-06 | 14.21 | -0.08 | 1.03E-21 | -0.07 | 2.50E-26 |
| rs1511243 | 8 | 76,230,943 | G | A | 98.51 | 0.11 | 0.01 | 83.60 | 0.08 | 2.20E-22 | 0.08 | 1.37E-23 |
| rs1533366 | 8 | 76,378,165 | T | G | 25.99 | -0.01 | 0.30 | 35.39 | -0.04 | 4.33E-12 | -0.04 | 1.37E-11 |
| rs12546444 | 8 | 106,358,620 | T | A | 11.31 | -0.08 | 1.10E-03 | 9.54 | -0.07 | 1.08E-11 | -0.07 | 4.90E-14 |
| rs13277568 | 8 | 116,679,547 | G | A | 44.05 | 0.00 | 0.75 | 35.79 | -0.04 | 2.23E-08 | -0.03 | 1.34E-06 |
| rs13267382 | 8 | 117,209,548 | G | A | 47.92 | -0.03 | 0.01 | 65.71 | -0.04 | 7.65E-12 | -0.04 | 3.69E-13 |
| rs62526620 | 8 | 120,862,186 | G | A | 2.58 | 0.11 | 6.29E-03 | 11.13 | 0.04 | 3.20E-05 | 0.04 | 3.47E-06 |
| rs35542655 | 8 | 124,563,705 | C | T | 18.25 | 0.03 | 0.07 | 15.21 | 0.06 | 1.12E-11 | 0.05 | 6.85E-12 |
| rs12541094 | 8 | 124,571,581 | A | G | 36.41 | 0.01 | 0.43 | 41.35 | 0.03 | 2.47E-08 | 0.03 | 7.97E-08 |
| rs7842619 | 8 | 124,739,913 | G | T | 24.21 | 0.02 | 0.20 | 39.86 | 0.04 | 3.04E-12 | 0.04 | 4.12E-12 |
| rs12550713 | 8 | 128,370,949 | G | C | 51.98 | 0.04 | 3.45E-04 | 43.24 | 0.10 | 1.46E-65 | 0.09 | 7.36E-64 |
| rs10096351 | 8 | 128,372,172 | G | A | 77.38 | 0.07 | 6.84E-06 | 56.76 | 0.11 | 3.77E-69 | 0.10 | 3.00E-72 |
| rs1016578 | 8 | 129,199,566 | A | G | 19.15 | -0.03 | 0.08 | 18.79 | 0.06 | 1.21E-15 | 0.04 | 5.29E-10 |
| rs7830152 | 8 | 143,669,254 | G | A | 82.94 | 0.00 | 0.95 | 33.10 | -0.02 | 3.31E-03 | -0.02 | 6.07E-03 |
| rs539723051 | 9 | 21,964,882 | C | CAAAA | 22.02 | 0.02 | 0.23 | 30.82 | 0.06 | 5.58E-22 | 0.06 | 3.16E-21 |
| rs17694493 | 9 | 22,041,998 | G | C | 1.39 | 0.09 | 0.05 | 13.02 | 0.05 | 1.34E-08 | 0.05 | 2.73E-09 |
| rs4880038 | 9 | 36,928,288 | C | T | 74.70 | 0.00 | 0.82 | 53.68 | 0.02 | 8.67E-05 | 0.02 | 4.75E-04 |
| rs665889 | 9 | 87,782,211 | C | T | 78.37 | 0.01 | 0.62 | 52.78 | 0.02 | 1.49E-03 | 0.02 | 1.87E-03 |
| rs10120432 | 9 | 98,362,587 | C | T | 34.42 | 0.01 | 0.55 | 8.55 | 0.04 | 5.16E-05 | 0.03 | 3.94E-04 |
| rs4742903 | 9 | 106,856,793 | C | G | 20.34 | 0.03 | 0.08 | 57.65 | 0.03 | 2.58E-08 | 0.03 | 5.87E-09 |
| rs60037937 | 9 | 110,303,808 | T | TAA | 43.25 | 0.03 | 8.98E-03 | 22.96 | 0.08 | 5.24E-26 | 0.07 | 2.42E-25 |
| rs10816625 | 9 | 110,837,073 | G | A | 38.19 | 0.08 | 2.15E-10 | 9.34 | 0.11 | 4.20E-20 | 0.10 | 3.96E-28 |
| rs13294895 | 9 | 110,837,176 | T | C | 2.08 | 0.04 | 0.44 | 18.89 | 0.06 | 5.00E-16 | 0.06 | 4.06E-16 |
| rs630965 | 9 | 110,885,479 | T | C | 91.57 | 0.05 | 0.01 | 63.12 | 0.10 | 1.95E-59 | 0.10 | 8.55E-60 |
| rs1895062 | 9 | 119,313,486 | G | A | 30.36 | 0.00 | 0.83 | 42.45 | -0.04 | 2.51E-12 | -0.04 | 1.33E-10 |
| rs3861871 | 9 | 129,424,719 | G | A | 57.94 | -0.06 | 1.40E-06 | 46.42 | -0.03 | 4.75E-08 | -0.04 | 2.16E-12 |
| rs550057 | 9 | 136,146,597 | T | C | 19.05 | 0.01 | 0.43 | 28.23 | 0.03 | 3.81E-07 | 0.03 | 9.55E-07 |
| rs55910451 | 10 | 5,794,652 | G | A | 15.97 | 0.02 | 0.26 | 20.08 | 0.04 | 6.93E-07 | 0.03 | 6.66E-07 |
| rs10796139 | 10 | 13,892,298 | A | G | 61.51 | 0.01 | 0.64 | 44.93 | 0.03 | 2.22E-05 | 0.02 | 5.46E-05 |
| rs7072776 | 10 | 22,032,942 | G | A | 94.54 | 0.01 | 0.79 | 71.37 | -0.06 | 2.51E-21 | -0.06 | 2.47E-20 |
| rs10764337 | 10 | 22,861,490 | C | A | 92.86 | -0.07 | 0.05 | 94.33 | 0.07 | 8.35E-09 | 0.06 | 2.37E-06 |

| | | | | | | | | | | | | |
|---|---|---|---|---|---|---|---|---|---|---|---|---|
| rs10995201 | 10 | 64,299,890 | G | A | 2.18 | -0.06 | 0.11 | 15.21 | -0.13 | 1.60E-49 | -0.12 | 1.71E-49 |
| rs6479868 | 10 | 64,819,996 | T | G | 16.77 | 0.03 | 0.04 | 19.38 | 0.03 | 1.44E-04 | 0.03 | 1.63E-05 |
| rs111833376 | 10 | 71,335,574 | T | C | 9.72 | 0.04 | 0.09 | 28.23 | -0.02 | 1.78E-03 | -0.02 | 0.01 |
| rs719338 | 10 | 80,851,257 | T | G | 66.47 | -0.06 | 4.28E-06 | 57.95 | -0.08 | 2.81E-37 | -0.07 | 1.86E-41 |
| rs4980029 | 10 | 80,886,726 | G | A | 46.43 | 0.06 | 1.85E-05 | 14.81 | 0.08 | 4.70E-22 | 0.07 | 1.12E-25 |
| rs140936696 | 10 | 95,292,187 | C | CAA | 62.80 | -0.01 | 0.32 | 82.50 | -0.04 | 8.37E-07 | -0.03 | 1.84E-06 |
| rs10885405 | 10 | 114,777,670 | T | C | 2.98 | 0.07 | 0.02 | 49.20 | 0.05 | 3.30E-14 | 0.05 | 3.53E-15 |
| rs12250948 | 10 | 115,128,491 | C | T | 24.70 | -0.05 | 4.18E-05 | 78.33 | -0.05 | 7.78E-14 | -0.05 | 1.55E-17 |
| rs9421410 | 10 | 123,095,209 | A | G | 40.48 | -0.04 | 1.39E-03 | 33.30 | -0.05 | 5.45E-13 | -0.05 | 3.31E-15 |
| rs45631580 | 10 | 123,340,107 | G | A | 11.31 | 0.04 | 0.04 | 7.16 | -0.13 | 5.35E-26 | -0.09 | 1.75E-15 |
| rs35054928 | 10 | 123,340,431 | G | GC | 55.06 | -0.18 | 3.94E-49 | 54.97 | -0.25 | 0.00E+00 | -0.23 | 3.87e-399 |
| rs6597981 | 11 | 803,017 | G | A | 27.58 | 0.03 | 0.02 | 49.80 | 0.05 | 4.65E-14 | 0.04 | 5.43E-15 |
| rs4980386 | 11 | 1,895,708 | A | C | 71.63 | -0.08 | 5.13E-08 | 39.96 | -0.08 | 4.89E-35 | -0.08 | 1.62E-41 |
| rs10832963 | 11 | 18,664,241 | G | T | 48.71 | 0.00 | 0.70 | 71.97 | 0.03 | 8.89E-06 | 0.03 | 5.27E-05 |
| rs4472923 | 11 | 42,844,441 | T | C | 22.82 | 0.02 | 0.17 | 34.79 | -0.01 | 0.07 | -0.01 | 0.24 |
| rs10838267 | 11 | 44,368,892 | A | G | 29.27 | 0.04 | 1.58E-03 | 51.69 | 0.03 | 4.51E-08 | 0.03 | 3.36E-10 |
| rs77047825 | 11 | 46,318,032 | G | C | 0.00 | -0.17 | 0.58 | 7.55 | -0.04 | 8.34E-04 | -0.04 | 7.72E-04 |
| rs12287832 | 11 | 65,553,492 | A | C | 11.11 | 0.03 | 0.10 | 18.49 | 0.05 | 1.45E-12 | 0.05 | 6.21E-13 |
| rs10896047 | 11 | 65,572,431 | A | G | 22.72 | -0.03 | 0.06 | 47.81 | -0.04 | 2.89E-13 | -0.04 | 7.99E-14 |
| rs35039974 | 11 | 69,328,130 | T | A | 23.81 | -0.02 | 0.14 | 18.29 | -0.07 | 1.15E-22 | -0.06 | 6.96E-21 |
| rs661204 | 11 | 69,330,983 | A | G | 0.99 | 0.25 | 1.77E-03 | 12.23 | 0.22 | 8.00E-137 | 0.22 | 6.55E-139 |
| rs78540526 | 11 | 69,331,418 | T | C | 0.50 | 0.26 | 0.02 | 6.76 | 0.28 | 2.09E-147 | 0.28 | 1.22E-148 |
| rs7125780 | 11 | 103,614,438 | G | T | 66.37 | -0.02 | 0.10 | 65.11 | 0.02 | 0.01 | 0.01 | 0.11 |
| rs199504893 | 11 | 108,267,402 | CA | C | 38.69 | 0.00 | 0.79 | 45.33 | 0.00 | 0.84 | 0.00 | 0.76 |
| rs610437 | 11 | 111,696,440 | C | T | 77.98 | -0.03 | 0.03 | 61.53 | -0.03 | 2.22E-07 | -0.03 | 2.07E-08 |
| rs625145 | 11 | 116,727,936 | T | A | 17.66 | -0.02 | 0.27 | 16.60 | -0.03 | 2.29E-04 | -0.03 | 1.58E-04 |
| rs7924772 | 11 | 120,233,626 | G | A | 24.21 | 0.03 | 0.08 | 38.87 | 0.02 | 6.93E-04 | 0.02 | 1.38E-04 |
| rs7121616 | 11 | 122,966,626 | G | A | 36.11 | -0.02 | 0.18 | 28.53 | -0.03 | 3.32E-05 | -0.03 | 1.75E-05 |
| rs7939702 | 11 | 129,243,417 | G | T | 96.63 | -0.04 | 0.41 | 84.99 | -0.05 | 1.87E-08 | -0.05 | 1.35E-08 |
| rs11822830 | 11 | 129,461,016 | G | A | 51.88 | 0.03 | 0.02 | 55.96 | 0.05 | 2.72E-14 | 0.04 | 4.37E-15 |
| rs797736 | 12 | 293,626 | G | A | 41.96 | 0.01 | 0.59 | 36.48 | 0.03 | 4.96E-05 | 0.02 | 1.24E-04 |
| rs12422552 | 12 | 14,413,931 | C | G | 26.98 | 0.07 | 6.95E-07 | 29.03 | 0.06 | 8.12E-18 | 0.06 | 3.91E-23 |
| rs788458 | 12 | 28,149,568 | T | C | 17.76 | -0.13 | 1.19E-16 | 10.64 | -0.15 | 5.89E-54 | -0.14 | 1.06E-68 |
| rs7297051 | 12 | 28,174,817 | T | C | 22.32 | -0.12 | 1.75E-16 | 23.26 | -0.12 | 2.38E-67 | -0.12 | 4.21E-82 |
| rs1027113 | 12 | 29,140,260 | A | G | 75.89 | 0.05 | 7.30E-04 | 92.84 | 0.07 | 7.67E-12 | 0.06 | 7.59E-14 |
| rs2277339 | 12 | 57,146,069 | G | T | 20.63 | -0.05 | 1.97E-03 | 11.13 | -0.04 | 1.29E-04 | -0.04 | 9.96E-07 |
| rs2870876 | 12 | 70,798,355 | T | A | 34.72 | -0.01 | 0.33 | 17.69 | 0.03 | 1.06E-04 | 0.02 | 5.00E-03 |
| rs111622698 | 12 | 83,064,195 | GA | G | 15.67 | 0.00 | 0.85 | 11.73 | 0.06 | 1.61E-07 | 0.04 | 6.94E-06 |
| rs10862899 | 12 | 85,004,551 | T | C | 11.90 | -0.02 | 0.25 | 49.70 | 0.03 | 1.19E-08 | 0.03 | 5.43E-07 |
| rs17356907 | 12 | 96,027,759 | G | A | 26.39 | -0.05 | 1.08E-03 | 29.32 | -0.09 | 8.11E-41 | -0.08 | 1.67E-41 |
| rs1061657 | 12 | 115,108,136 | C | T | 41.67 | 0.01 | 0.49 | 28.13 | 0.04 | 2.52E-10 | 0.04 | 4.11E-09 |

| | | | | | | | | | | | | |
|---|---|---|---|---|---|---|---|---|---|---|---|---|
| rs11067551 | 12 | 115,796,577 | G | A | 29.07 | -0.05 | 1.43E-04 | 20.28 | -0.04 | 9.39E-08 | -0.04 | 6.86E-11 |
| rs2454399 | 12 | 115,835,836 | C | T | 25.99 | -0.10 | 5.53E-14 | 41.35 | -0.08 | 1.13E-42 | -0.09 | 9.01E-55 |
| rs2464195 | 12 | 121,435,475 | A | G | 48.02 | -0.03 | 6.08E-03 | 37.77 | -0.02 | 1.88E-03 | -0.02 | 5.48E-05 |
| rs9315973 | 13 | 43,501,356 | G | A | 61.61 | -0.01 | 0.46 | 80.72 | 0.04 | 1.39E-05 | 0.02 | 8.02E-04 |
| rs12870942 | 13 | 73,806,982 | C | T | 23.41 | 0.05 | 4.22E-04 | 30.42 | 0.04 | 3.79E-11 | 0.04 | 7.60E-14 |
| rs2181965 | 13 | 73,960,952 | G | A | 99.50 | -0.02 | 0.84 | 77.04 | 0.04 | 6.50E-10 | 0.04 | 8.00E-10 |
| rs34914085 | 14 | 37,128,564 | A | C | 31.65 | -0.07 | 7.18E-07 | 21.57 | -0.07 | 2.66E-22 | -0.07 | 1.18E-27 |
| rs2253012 | 14 | 37,228,504 | T | C | 10.32 | 0.05 | 0.01 | 43.34 | 0.04 | 5.04E-10 | 0.04 | 2.27E-11 |
| rs2588809 | 14 | 68,660,428 | C | T | 97.22 | -0.08 | 0.02 | 81.21 | -0.06 | 1.36E-14 | -0.06 | 1.23E-15 |
| rs11624333 | 14 | 68,979,835 | C | T | 5.56 | -0.01 | 0.71 | 23.66 | -0.10 | 1.55E-43 | -0.09 | 2.03E-40 |
| rs11341843 | 14 | 91,751,788 | T | TC | 49.31 | 0.03 | 0.03 | 67.69 | 0.05 | 1.57E-13 | 0.04 | 4.99E-14 |
| rs941764 | 14 | 91,841,069 | G | A | 13.69 | 0.03 | 0.10 | 35.29 | 0.05 | 4.11E-15 | 0.05 | 1.87E-15 |
| rs4983544 | 14 | 105,213,978 | G | T | 65.97 | 0.02 | 0.17 | 44.63 | 0.04 | 2.04E-09 | 0.03 | 2.06E-09 |
| rs4774565 | 15 | 50,694,306 | G | A | 51.59 | 0.01 | 0.56 | 33.80 | -0.03 | 2.76E-06 | -0.03 | 2.65E-05 |
| rs8042593 | 15 | 66,630,569 | A | G | 81.15 | 0.00 | 0.92 | 61.53 | -0.03 | 1.61E-05 | -0.02 | 5.92E-05 |
| rs35874463 | 15 | 67,457,698 | G | A | 0.00 | 0.31 | 0.37 | 5.27 | 0.07 | 2.59E-07 | 0.07 | 2.19E-07 |
| rs8035987 | 15 | 75,750,383 | C | T | 37.90 | -0.05 | 1.76E-04 | 28.93 | -0.03 | 5.94E-07 | -0.04 | 5.98E-10 |
| rs2290202 | 15 | 91,512,267 | T | G | 51.29 | -0.07 | 3.34E-09 | 14.02 | -0.08 | 1.04E-17 | -0.07 | 2.36E-25 |
| rs144767203 | 15 | 100,905,819 | C | A | 22.92 | -0.03 | 0.15 | 9.54 | -0.05 | 4.22E-06 | -0.04 | 2.64E-06 |
| rs11076805 | 16 | 4,106,788 | A | C | 11.41 | -0.03 | 0.26 | 25.75 | -0.03 | 1.14E-04 | -0.03 | 5.77E-05 |
| rs12709163 | 16 | 6,963,972 | G | C | 88.79 | -0.04 | 0.09 | 79.03 | 0.01 | 0.14 | 0.01 | 0.43 |
| rs34872983 | 16 | 10,706,580 | A | G | 23.81 | -0.02 | 0.15 | 5.07 | -0.07 | 1.85E-07 | -0.05 | 6.32E-07 |
| rs75753503 | 16 | 23,007,047 | T | G | 3.27 | -0.01 | 0.82 | 2.49 | 0.07 | 8.57E-04 | 0.05 | 4.95E-03 |
| rs35668161 | 16 | 52,538,825 | A | C | 18.75 | 0.20 | 1.64E-37 | 27.24 | 0.21 | 4.88E-212 | 0.21 | 1.55E-247 |
| rs4784227 | 16 | 52,599,188 | T | C | 25.50 | 0.21 | 1.18E-51 | 25.45 | 0.21 | 5.65E-214 | 0.21 | 1.34E-263 |
| rs55872725 | 16 | 53,809,123 | T | C | 16.57 | -0.07 | 1.52E-05 | 43.24 | -0.06 | 2.20E-22 | -0.06 | 2.09E-26 |
| rs6499648 | 16 | 53,861,139 | T | C | 67.66 | -0.03 | 0.03 | 78.53 | -0.04 | 2.48E-09 | -0.04 | 3.34E-10 |
| rs7184573 | 16 | 53,861,592 | A | G | 18.85 | -0.01 | 0.37 | 39.66 | -0.05 | 3.20E-14 | -0.04 | 1.33E-13 |
| rs28539243 | 16 | 54,682,064 | A | G | 58.04 | 0.03 | 8.55E-03 | 47.91 | 0.05 | 1.52E-14 | 0.04 | 7.63E-16 |
| rs7500067 | 16 | 80,648,296 | G | A | 25.99 | 0.04 | 4.89E-03 | 24.65 | 0.08 | 1.68E-30 | 0.07 | 2.86E-30 |
| rs9931038 | 16 | 85,145,977 | C | T | 11.61 | 0.00 | 0.94 | 46.12 | -0.01 | 0.08 | -0.01 | 0.09 |
| rs12449271 | 16 | 87,086,492 | C | T | 20.14 | -0.03 | 0.06 | 26.44 | -0.04 | 9.24E-10 | -0.04 | 1.96E-10 |
| rs79461387 | 17 | 29,168,077 | T | G | 9.52 | -0.06 | 0.06 | 27.63 | -0.04 | 1.22E-09 | -0.04 | 2.22E-10 |
| rs545502941 | 17 | 43,212,339 | CT | C | 9.72 | -0.02 | 0.34 | 22.76 | 0.03 | 7.28E-05 | 0.02 | 8.11E-04 |
| rs559816018 | 17 | 44,283,858 | A | G | 0.10 | -0.12 | 0.54 | 19.48 | -0.05 | 1.68E-11 | -0.05 | 1.47E-11 |
| rs2787486 | 17 | 53,209,774 | C | A | 27.98 | -0.08 | 4.02E-09 | 28.63 | -0.07 | 1.46E-27 | -0.07 | 4.99E-35 |
| rs11652463 | 17 | 70,405,095 | G | C | 74.80 | -0.01 | 0.30 | 31.01 | -0.04 | 4.19E-08 | -0.03 | 8.09E-08 |
| rs745570 | 17 | 77,781,725 | G | A | 41.27 | -0.04 | 4.48E-03 | 49.50 | -0.04 | 1.56E-10 | -0.04 | 2.58E-12 |
| rs206435 | 18 | 10,354,649 | C | A | 49.70 | 0.00 | 0.85 | 50.00 | 0.00 | 0.99 | 0.00 | 0.93 |
| rs16976596 | 18 | 11,696,613 | T | C | 2.98 | -0.05 | 0.46 | 14.91 | -0.03 | 3.34E-03 | -0.03 | 2.59E-03 |
| rs11665269 | 18 | 20,634,253 | T | C | 48.61 | 0.01 | 0.66 | 62.43 | -0.04 | 2.35E-08 | -0.03 | 2.19E-06 |

| | | | | | | | | | | | | |
|---|---|---|---|---|---|---|---|---|---|---|---|---|
| rs1111207 | 18 | 24,125,857 | C | T | 66.37 | 0.02 | 0.09 | 43.14 | 0.03 | 6.44E-08 | 0.03 | 2.00E-08 |
| rs527616 | 18 | 24,337,424 | G | C | 74.21 | 0.04 | 3.69E-03 | 60.44 | 0.05 | 1.68E-16 | 0.05 | 3.41E-18 |
| rs35369219 | 18 | 24,518,050 | A | AT | 49.21 | -0.06 | 1.87E-06 | 27.34 | -0.06 | 1.67E-20 | -0.06 | 1.75E-25 |
| rs8092192 | 18 | 25,407,513 | G | C | 45.44 | 0.01 | 0.48 | 70.87 | 0.03 | 7.84E-06 | 0.02 | 2.05E-05 |
| rs72931898 | 18 | 29,981,526 | A | G | 1.09 | -0.07 | 0.31 | 4.08 | -0.10 | 9.68E-13 | -0.10 | 6.24E-13 |
| rs9954058 | 18 | 42,411,803 | C | G | 13.49 | -0.08 | 1.06E-05 | 7.26 | -0.08 | 1.30E-12 | -0.08 | 6.83E-17 |
| rs9952980 | 18 | 42,888,797 | C | T | 30.95 | -0.09 | 1.96E-12 | 34.00 | -0.05 | 1.08E-14 | -0.06 | 8.31E-24 |
| rs56069439 | 19 | 17,393,925 | A | C | 0.20 | -0.05 | 0.67 | 26.04 | 0.04 | 8.98E-09 | 0.04 | 1.10E-08 |
| rs10164323 | 19 | 18,569,492 | T | C | 21.63 | -0.05 | 2.54E-04 | 35.09 | -0.07 | 2.08E-27 | -0.07 | 5.53E-30 |
| rs140702307 | 19 | 19,517,054 | CGGGCG | C | 40.67 | 0.04 | 9.18E-03 | 34.00 | 0.04 | 4.14E-10 | 0.04 | 1.32E-11 |
| rs56681946 | 19 | 44,283,031 | C | T | 15.08 | -0.01 | 0.58 | 34.19 | 0.06 | 2.34E-22 | 0.05 | 1.32E-19 |
| rs4399645 | 19 | 46,166,073 | C | T | 40.18 | -0.03 | 0.06 | 60.64 | -0.04 | 3.37E-08 | -0.03 | 6.26E-09 |
| rs1172821 | 19 | 55,816,678 | T | C | 9.82 | -0.02 | 0.35 | 35.59 | -0.03 | 3.52E-06 | -0.03 | 2.29E-06 |
| rs16991615 | 20 | 5,948,227 | A | G | 0.00 | 0.13 | 0.75 | 6.66 | 0.08 | 7.48E-10 | 0.08 | 7.18E-10 |
| rs6065254 | 20 | 39,248,265 | A | G | 39.48 | -0.01 | 0.32 | 41.05 | -0.03 | 1.99E-05 | -0.02 | 1.99E-05 |
| rs6030585 | 20 | 41,613,706 | G | C | 84.52 | -0.02 | 0.18 | 80.42 | 0.02 | 0.04 | 0.01 | 0.16 |
| rs13039563 | 20 | 52,296,849 | A | G | 32.04 | 0.04 | 4.88E-03 | 24.25 | 0.04 | 3.05E-09 | 0.04 | 5.48E-11 |
| rs2822999 | 21 | 16,364,756 | G | T | 15.18 | 0.00 | 1.00 | 16.90 | 0.06 | 2.86E-12 | 0.05 | 6.88E-10 |
| rs2823130 | 21 | 16,566,350 | G | A | 12.90 | 0.07 | 1.05E-04 | 9.44 | 0.07 | 3.52E-10 | 0.07 | 1.66E-13 |
| rs2403907 | 21 | 16,574,455 | A | C | 11.61 | -0.04 | 0.03 | 29.22 | -0.08 | 1.38E-33 | -0.07 | 7.99E-34 |
| rs4818836 | 21 | 47,762,932 | A | G | 0.10 | 0.24 | 7.59E-03 | 3.78 | 0.08 | 8.81E-07 | 0.09 | 1.02E-07 |
| rs9798754 | 22 | 19,766,137 | T | C | 58.63 | -0.04 | 2.53E-03 | 36.38 | -0.03 | 6.59E-08 | -0.04 | 6.64E-10 |
| rs5997390 | 22 | 29,135,543 | A | G | 6.35 | 0.07 | 1.02E-03 | 7.95 | 0.07 | 2.42E-12 | 0.07 | 1.02E-14 |
| rs536920426 | 22 | 38,583,315 | AAAAGAAAG | AAAAG | 23.51 | -0.01 | 0.61 | 29.42 | -0.05 | 4.32E-13 | -0.05 | 3.33E-12 |
| rs5750715 | 22 | 39,343,916 | A | T | 47.12 | 0.06 | 4.29E-07 | 27.44 | 0.04 | 2.05E-10 | 0.05 | 1.16E-15 |
| rs66987842 | 22 | 40,904,707 | C | CT | 23.91 | 0.06 | 2.83E-06 | 10.44 | 0.12 | 7.18E-36 | 0.10 | 3.30E-38 |
| rs112855987 | 22 | 45,319,953 | A | G | 29.76 | -0.03 | 0.04 | 41.75 | -0.01 | 0.08 | -0.01 | 0.02 |

[a] The 330-SNP European PRS for breast cancer reported by Zhang et al. *Nat Genet*. 2020. Of the 330 SNPs, 263 SNPs were available in our validation and prospective test sets. Based on these 263 SNPs, $PRS_{263\text{-}ASN}$, $PRS_{263\text{-}EUR}$ and $PRS_{263\text{-}META}$ were derived using weights from our training set, BCAC-European data, and meta-analyses of these two datasets respectively.
[b] BCAC-European data from Zhang et al. *Nat Genet*. 2020.

### eTable 5. Associations of the 111 SNPs in $PRS_{111}$ with breast cancer risk in our ABCC training set and BCAC-European data

| RSID | Chr | Position (GRCh37) | Effect allele | Non-effect allele | ABCC training set | | | BCAC-European data | | |
|---|---|---|---|---|---|---|---|---|---|---|
| | | | | | Effect allele frequency (%) | Effect size | *P* | Effect allele frequency (%) | Effect size [c] | *P* [c] |
| **57 SNPs selected by fine-mapping at COJO-*P*<$10^{-5}$ using our training set [a]** | | | | | | | | | | |
| rs4846235 | 1 | 10,632,235 | T | C | 20.24 | -0.080 | 2.13E-07 | 31.11 | -0.06 | 6.46E-21 |
| rs7529564 | 1 | 156,189,793 | T | C | 86.21 | -0.072 | 7.54E-06 | 63.82 | -0.03 | 1.18E-06 |
| rs67087079 | 1 | 203,850,783 | A | G | 24.60 | 0.072 | 1.89E-07 | 11.43 | 0.05 | 1.01E-09 |
| rs12127615 | 1 | 88,181,633 | A | G | 22.32 | -0.068 | 7.36E-06 | 59.24 | -0.01 | 0.03 |
| rs4848601 | 2 | 121,243,011 | C | G | 79.07 | 0.086 | 3.23E-08 | 89.76 | 0.10 | 2.17E-22 |
| rs10931936 | 2 | 202,143,928 | T | C | 30.36 | 0.103 | 6.24E-16 | 28.33 | 0.06 | 2.72E-17 |
| rs57481445 | 2 | 218,296,374 | A | G | 41.77 | -0.064 | 8.68E-08 | 72.56 | -0.07 | 5.57E-26 |
| rs34197427 | 3 | 27,532,310 | A | G | 14.19 | 0.096 | 4.70E-08 | 37.57 | 0.05 | 1.01E-14 |
| rs6440015 | 3 | 141,336,351 | T | G | 29.56 | 0.061 | 2.12E-06 | 37.97 | 0.03 | 2.46E-07 |
| rs73010941 | 3 | 150,474,477 | T | C | 56.15 | 0.103 | 4.19E-16 | 97.22 | 0.05 | 7.33E-03 |
| rs2945330 | 4 | 48,606,526 | A | C | 62.80 | 0.066 | 1.51E-07 | 48.91 | 0.02 | 6.79E-04 |
| rs9884717 | 4 | 175,833,091 | A | G | 76.59 | 0.081 | 1.04E-08 | 89.17 | 0.10 | 6.25E-27 |
| rs4339357 | 5 | 44,670,741 | T | C | 45.24 | -0.104 | 4.23E-18 | 59.15 | -0.09 | 3.05E-53 |
| rs112776581 | 5 | 56,054,333 | T | TA | 11.01 | 0.194 | 1.11E-20 | 4.87 | 0.20 | 5.02E-55 |
| rs6860948 | 5 | 90,696,297 | T | G | 49.40 | 0.071 | 6.53E-09 | 85.79 | 0.06 | 1.35E-12 |
| rs17715065 | 5 | 158,261,163 | T | C | 38.19 | -0.068 | 4.37E-08 | 49.30 | -0.07 | 4.93E-29 |
| rs2444832 | 6 | 81,339,849 | A | T | 31.85 | 0.057 | 5.12E-06 | 38.97 | 0.02 | 2.26E-03 |
| rs9444166 | 6 | 85,088,902 | A | C | 79.96 | 0.072 | 2.12E-06 | 68.39 | 0.03 | 9.02E-05 |
| rs4897114 | 6 | 149,607,978 | A | G | 42.56 | -0.084 | 1.41E-12 | 16.40 | -0.04 | 1.71E-06 |
| rs7763637 | 6 | 151,949,312 | A | G | 34.03 | 0.215 | 1.99E-62 | 29.82 | 0.09 | 1.30E-44 |
| rs862346 | 6 | 152,016,369 | A | T | 87.70 | -0.083 | 4.38E-07 | 55.27 | -0.05 | 1.47E-14 |
| rs79388591 | 6 | 152,355,649 | G | GT | 67.46 | -0.096 | 1.52E-13 | 92.15 | -0.04 | 3.22E-04 |
| rs9397082 | 6 | 152,430,638 | T | C | 36.11 | 0.063 | 2.91E-07 | 27.63 | 0.05 | 3.69E-12 |
| rs2172905 | 6 | 170,334,502 | T | C | 78.77 | -0.067 | 4.37E-06 | 28.83 | -0.03 | 8.31E-06 |
| rs17164125 | 7 | 91,417,796 | T | C | 61.61 | 0.062 | 5.99E-07 | 90.85 | 0.03 | 2.19E-03 |
| rs13235624 | 7 | 139,943,267 | T | C | 46.43 | 0.057 | 4.47E-06 | 43.24 | 0.05 | 2.81E-17 |
| rs4732987 | 8 | 29,494,941 | T | C | 51.69 | 0.062 | 4.65E-07 | 60.54 | 0.03 | 2.90E-07 |
| rs146992477 | 8 | 36,842,055 | T | TTCTTTCTTTC | 68.95 | 0.069 | 1.64E-07 | 86.28 | 0.07 | 4.28E-18 |
| rs34302508 | 8 | 102,654,384 | CT | C | 91.07 | 0.112 | 1.71E-07 | 96.82 | 0.04 | 0.02 |
| rs2392780 | 8 | 128,388,025 | A | G | 73.81 | 0.073 | 1.84E-07 | 58.95 | 0.10 | 5.92E-63 |
| rs1333035 | 9 | 22,044,059 | A | G | 81.94 | -0.089 | 6.96E-10 | 90.36 | -0.03 | 8.17E-04 |
| rs10816625 | 9 | 110,837,073 | A | G | 61.81 | -0.079 | 2.21E-10 | 90.66 | -0.11 | 4.20E-20 |
| rs10760444 | 9 | 129,396,434 | A | G | 43.25 | -0.069 | 1.13E-08 | 55.17 | -0.03 | 9.63E-09 |
| rs78053936 | 10 | 64,300,331 | A | C | 76.98 | 0.110 | 4.54E-11 | 98.01 | 0.06 | 4.55E-04 |
| rs2252004 | 10 | 122,844,709 | A | C | 31.65 | -0.070 | 2.63E-07 | 8.75 | -0.03 | 1.72E-03 |
| rs2248051 | 10 | 122,854,749 | T | G | 37.50 | -0.065 | 5.19E-07 | 8.75 | -0.03 | 1.97E-03 |
| rs7913903 | 10 | 123,095,094 | A | G | 32.54 | -0.067 | 4.31E-07 | 24.35 | -0.03 | 6.19E-06 |
| rs2912778 | 10 | 123,338,654 | A | G | 43.65 | -0.198 | 5.23E-62 | 47.42 | -0.21 | 1.95E-263 |

| | | | | | | | | | | |
|---|---|---|---|---|---|---|---|---|---|---|
| rs509239 | 11 | 1,885,117 | A | T | 23.41 | 0.095 | 6.17E-12 | 35.29 | 0.04 | 4.39E-10 |
| rs1873872 | 11 | 129,473,993 | A | T | 65.08 | -0.057 | 2.28E-06 | 47.71 | -0.04 | 2.30E-12 |
| rs12422552 | 12 | 14,413,931 | C | G | 26.98 | 0.068 | 7.47E-07 | 29.03 | 0.06 | 8.12E-18 |
| rs1314084 | 12 | 28,140,277 | T | C | 82.54 | 0.136 | 1.84E-17 | 86.78 | 0.12 | 1.36E-42 |
| rs833734 | 12 | 103,044,493 | A | G | 6.15 | -0.103 | 9.02E-06 | 3.18 | -0.08 | 2.49E-05 |
| rs11067567 | 12 | 115,828,256 | T | C | 23.61 | -0.109 | 8.28E-16 | 34.49 | -0.08 | 1.59E-34 |
| rs12895715 | 14 | 37,113,093 | A | G | 67.66 | 0.070 | 1.35E-07 | 78.13 | 0.07 | 6.96E-22 |
| rs8037137 | 15 | 91,506,637 | T | C | 48.12 | 0.073 | 1.98E-09 | 86.08 | 0.08 | 1.50E-17 |
| rs112149573 | 16 | 52,581,245 | T | G | 24.70 | 0.213 | 3.95E-52 | 24.95 | 0.21 | 1.08E-208 |
| rs17817964 | 16 | 53,828,066 | T | C | 18.35 | -0.075 | 1.35E-06 | 41.15 | -0.06 | 9.09E-20 |
| rs3893264 | 16 | 54,683,802 | T | C | 84.72 | -0.089 | 9.54E-06 | 81.51 | -0.05 | 9.53E-12 |
| rs149288672 | 16 | 71,916,281 | CAG | C | 75.20 | 0.070 | 1.05E-06 | 85.98 | 0.04 | 6.89E-05 |
| rs244373 | 17 | 53,184,949 | T | C | 28.67 | -0.084 | 8.32E-10 | 28.43 | -0.07 | 2.17E-27 |
| rs2307561 | 18 | 24,503,506 | A | AAGTGTT | 49.40 | -0.062 | 1.48E-06 | 27.04 | -0.07 | 1.68E-21 |
| rs10502843 | 18 | 42,378,282 | A | G | 14.48 | -0.084 | 4.70E-06 | 6.46 | -0.08 | 1.61E-10 |
| rs12455117 | 18 | 42,884,026 | A | T | 70.34 | 0.094 | 2.55E-13 | 66.90 | 0.05 | 1.52E-12 |
| rs2823126 | 21 | 16,561,704 | A | G | 30.75 | -0.091 | 1.32E-10 | 2.19 | -0.05 | 0.02 |
| rs6001335 | 22 | 39,345,966 | C | G | 47.22 | 0.064 | 4.14E-07 | 27.24 | 0.04 | 2.81E-10 |
| rs141580207 | 22 | 40,917,540 | T | TCA | 75.40 | -0.067 | 7.50E-07 | 91.05 | -0.12 | 4.22E-33 |
| **54 index SNPs in GWAS-identified loci that were not eligible for fine-mapping [b]** | | | | | | | | | | |
| rs72906468 | 1 | 17,772,093 | A | T | 66.87 | 0.054 | 8.00E-05 | 80.02 | 0.03 | 1.82E-06 |
| rs2992756 | 1 | 18,807,339 | T | C | 14.29 | 0.045 | 6.83E-03 | 51.19 | 0.05 | 2.67E-17 |
| rs3790585 | 1 | 46,023,356 | A | T | 67.76 | 0.032 | 0.02 | 85.88 | 0.04 | 8.87E-07 |
| rs11249433 | 1 | 121,280,613 | A | G | 97.02 | -0.089 | 0.01 | 56.86 | -0.10 | 1.21E-57 |
| rs12710696 | 2 | 19,320,803 | T | C | 30.75 | 0.037 | 4.28E-03 | 34.19 | 0.04 | 2.68E-09 |
| rs71801447 | 2 | 111,925,731 | CTTATGTT | C | 89.48 | -0.051 | 5.04E-03 | 92.94 | -0.06 | 4.23E-07 |
| rs6762644 | 3 | 4,742,276 | A | G | 93.45 | -0.053 | 0.01 | 64.61 | -0.05 | 1.15E-18 |
| rs12493607 | 3 | 30,682,939 | C | G | 70.63 | 0.039 | 1.68E-03 | 33.70 | 0.05 | 2.72E-14 |
| rs6796502 | 3 | 46,866,866 | A | G | 13.79 | -0.035 | 0.03 | 9.05 | -0.08 | 1.23E-15 |
| rs1053338 | 3 | 63,967,900 | A | G | 85.22 | -0.036 | 0.03 | 85.29 | -0.06 | 1.42E-11 |
| rs58058861 | 3 | 172,285,237 | A | G | 32.74 | 0.031 | 0.03 | 17.89 | 0.05 | 2.40E-10 |
| rs10069690 | 5 | 1,279,790 | T | C | 16.87 | 0.061 | 3.52E-03 | 27.63 | 0.06 | 1.76E-18 |
| rs3215401 | 5 | 1,296,255 | A | AG | 61.61 | 0.035 | 0.02 | 71.17 | 0.07 | 1.13E-23 |
| rs6555134 | 5 | 2,776,483 | T | C | 23.21 | -0.034 | 0.02 | 56.96 | -0.03 | 5.13E-07 |
| rs204247 | 6 | 13,722,523 | A | G | 38.49 | -0.043 | 3.63E-04 | 57.75 | -0.05 | 1.56E-14 |
| rs7765429 | 6 | 21,904,169 | T | C | 83.04 | -0.056 | 9.91E-03 | 49.11 | -0.04 | 1.65E-09 |
| rs17529111 | 6 | 82,128,386 | T | C | 78.17 | -0.034 | 0.02 | 77.73 | -0.05 | 2.75E-10 |
| rs17268829 | 7 | 94,113,799 | T | C | 71.53 | -0.051 | 5.55E-04 | 71.17 | -0.05 | 2.58E-13 |
| rs4593472 | 7 | 130,667,121 | T | C | 16.27 | -0.044 | 6.31E-03 | 33.20 | -0.04 | 4.93E-11 |
| rs144145984 | 8 | 23,644,003 | CT | C | 45.04 | -0.041 | 8.60E-04 | 57.85 | -0.03 | 1.46E-05 |
| rs6472903 | 8 | 76,230,301 | T | G | 96.83 | 0.129 | 9.42E-05 | 83.60 | 0.08 | 2.48E-22 |
| rs2849506 | 8 | 101,329,134 | C | G | 46.83 | -0.042 | 4.10E-04 | 38.97 | -0.03 | 1.87E-05 |
| rs12546444 | 8 | 106,358,620 | A | T | 88.69 | 0.078 | 1.10E-03 | 90.46 | 0.07 | 1.08E-11 |
| rs13267382 | 8 | 117,209,548 | A | G | 52.08 | 0.034 | 0.01 | 34.29 | 0.04 | 7.65E-12 |
| rs142360995 | 8 | 118,205,719 | A | G | 7.64 | 0.077 | 2.08E-03 | 19.98 | 0.03 | 1.68E-04 |

| | | | | | | | | | | |
|---|---|---|---|---|---|---|---|---|---|---|
| rs58847541 | 8 | 124,610,166 | A | G | 18.75 | 0.033 | 0.04 | 15.31 | 0.06 | 1.91E-13 |
| rs10759243 | 9 | 110,306,115 | A | C | 45.04 | 0.036 | 3.22E-03 | 30.82 | 0.06 | 3.08E-17 |
| rs7904519 | 10 | 114,773,927 | A | G | 96.53 | -0.078 | 0.01 | 50.50 | -0.04 | 8.99E-14 |
| rs2901157 | 10 | 119,262,365 | A | G | 77.08 | 0.055 | 8.46E-05 | 87.38 | 0.04 | 1.40E-05 |
| rs6597981 | 11 | 803,017 | A | G | 72.42 | -0.031 | 0.02 | 50.20 | -0.05 | 4.65E-14 |
| rs10838267 | 11 | 44,368,892 | A | G | 29.27 | 0.043 | 1.58E-03 | 51.69 | 0.03 | 4.51E-08 |
| rs1027113 | 12 | 29,140,260 | A | G | 75.89 | 0.048 | 7.30E-04 | 92.84 | 0.07 | 7.67E-12 |
| rs78588049 | 12 | 69,180,907 | A | ATTTT | 17.76 | -0.053 | 2.92E-03 | 20.87 | -0.03 | 1.22E-05 |
| rs17356907 | 12 | 96,027,759 | A | G | 73.61 | 0.046 | 1.08E-03 | 70.68 | 0.09 | 8.11E-41 |
| rs2464195 | 12 | 121,435,475 | A | G | 48.02 | -0.032 | 6.08E-03 | 37.77 | -0.02 | 1.88E-03 |
| rs9316500 | 13 | 51,094,114 | T | G | 35.22 | 0.046 | 1.92E-04 | 72.76 | 0.03 | 4.37E-06 |
| rs2588809 | 14 | 68,660,428 | T | C | 2.78 | 0.084 | 0.02 | 18.79 | 0.06 | 1.36E-14 |
| rs75004998 | 14 | 77,517,786 | A | G | 46.63 | -0.036 | 3.31E-03 | 34.59 | -0.03 | 3.40E-06 |
| rs11627032 | 14 | 93,104,072 | T | C | 74.01 | 0.027 | 0.04 | 73.46 | 0.05 | 2.20E-11 |
| rs8027365 | 15 | 75,808,740 | A | C | 65.38 | 0.042 | 6.33E-04 | 69.78 | 0.03 | 3.91E-07 |
| rs2432539 | 16 | 56,420,987 | A | G | 38.99 | 0.038 | 2.59E-03 | 41.65 | 0.03 | 5.08E-07 |
| rs4496150 | 16 | 87,085,237 | A | C | 45.73 | -0.031 | 0.01 | 26.04 | -0.04 | 1.25E-09 |
| rs146699004 | 17 | 29,230,520 | G | GGT | 9.72 | -0.058 | 0.04 | 28.63 | -0.04 | 6.52E-09 |
| rs745570 | 17 | 77,781,725 | A | G | 58.73 | 0.035 | 4.48E-03 | 50.50 | 0.04 | 1.56E-10 |
| rs78269692 | 19 | 13,158,277 | T | C | 100.00 | -1.101 | 0.01 | 95.92 | -0.09 | 1.14E-09 |
| rs2594714 | 19 | 13,954,571 | A | G | 25.69 | -0.031 | 0.03 | 24.35 | -0.04 | 2.92E-08 |
| rs4808801 | 19 | 18,571,141 | A | G | 78.37 | 0.050 | 2.45E-04 | 65.51 | 0.07 | 6.64E-29 |
| rs2965183 | 19 | 19,545,696 | A | G | 32.04 | 0.039 | 3.15E-03 | 33.70 | 0.04 | 3.58E-10 |
| rs113701136 | 19 | 30,277,729 | T | C | 22.02 | 0.040 | 6.34E-03 | 28.13 | 0.02 | 2.08E-03 |
| rs71338792 | 19 | 46,183,031 | A | AT | 78.57 | -0.060 | 2.29E-04 | 78.23 | -0.04 | 1.28E-08 |
| rs12481286 | 20 | 52,287,610 | T | G | 32.64 | 0.042 | 2.34E-03 | 24.06 | 0.04 | 6.10E-09 |
| rs9808759 | 21 | 47,780,223 | T | C | 18.55 | 0.029 | 0.05 | 7.16 | 0.07 | 5.84E-09 |
| rs35418111 | 21 | 47,856,670 | A | G | 19.25 | 0.037 | 0.01 | 6.96 | 0.07 | 2.10E-08 |
| rs34331122 | 22 | 19,762,428 | CTT | C | 57.24 | -0.057 | 6.62E-05 | 43.34 | -0.03 | 6.56E-08 |

[a] 66 SNPs were selected by fine-mapping at COJO-*P*<$10^{-5}$ and 57 of them showed consistent association patterns with breast cancer risk at *P*<0.05 in BCAC-European data. Effect sizes and *P* values of these 57 SNPs were derived from a joint analysis of all SNPs selected by fine-mapping within each loci.

[b] A total of 54 SNPs in loci that were ineligible for fine-mapping showed *P*<0.05 in our training set. Effect sizes and *P* values of these 57 SNPs were from our training set.

[c] BCAC-European data from Zhang et al. *Nat Genet*. 2020.

### eTable 6. Risk stratification performance of PRSs, the NgRS, and IRSs in the prospective test set

| Score | Percentage of women with an OR>3 compared with average risk group | | Percentage of women with an OR>2 compared with average risk group | |
|---|---|---|---|---|
| | Percentage | OR (95% CI) [a] | Percentage | OR (95% CI) [a] |
| NgRS [b] | NA | NA | 6.9% | 2.00 (1.22-3.28) |
| $PRS_{111}$ [c] | 11.3% | 3.02 (1.99-4.63) | 37.6% | 2.00 (1.42-2.86) |
| $IRS_{111}$ [c] | 14.0% | 3.00 (2.02-4.51) | 38.7% | 2.00 (1.41-2.86) |
| $PRS_{263\text{-}META}$ [d] | 2.9% | 3.08 (1.71-5.69) | 8.6% | 2.02 (1.32-3.11) |
| $IRS_{263\text{-}META}$ [d] | 4.4% | 3.00 (1.80-5.08) | 13.4% | 2.01 (1.36-2.97) |

PRS, polygenic risk score; NgRS, nongenetic risk score; IRS, integrated risk score; OR, odds ratio; CI, confidence interval.

[a] ORs and 95% CIs of PRS/NgRS/IRS percentile groups compared with the average risk group (40%-60% percentiles) were estimated using logistic regression.

[b] Based on weighted six nongenetic risk factors and an interaction item.

[c] $PRS_{111}$: the best PRS in the present study. $IRS_{111}$: the combination of $PRS_{111}$ and the NgRS.

[d] $PRS_{263\text{-}META}$ was derived based on the 330-SNP European PRS reported by Zhang et al. *Nat Genet*. 2020. $PRS_{263\text{-}META}$ was the combination of $PRS_{263\text{-}EUR}$ and the NgRS.

### eTable 7. Association with breast cancer risk for $PRS_{111}$, $IRS_{111}$, $PRS_{263\text{-}META}$ and $IRS_{263\text{-}META}$ in 10,207 Chinese women [a]

| Percentiles | $PRS_{111}$ [b] | | $IRS_{111}$ [b] | | $PRS_{263\text{-}META}$ [c] | | $IRS_{263\text{-}META}$ [c] | |
|---|---|---|---|---|---|---|---|---|
| | OR (95% CI) [d] | *P* [d] | OR (95% CI) [d] | *P* [d] | OR (95% CI) [d] | *P* [d] | OR (95% CI) [d] | *P* [d] |
| <5 | 0.30 (0.24-0.39) | 5.124E-20 | 0.27 (0.21-0.35) | 3.83E-22 | 0.44 (0.35-0.56) | 1.33E-11 | 0.36 (0.27-0.47) | 1.14E-13 |
| 5-10 | 0.54 (0.43-0.67) | 2.81E-08 | 0.48 (0.38-0.60) | 2.03E-10 | 0.51 (0.40-0.63) | 4.45E-09 | 0.45 (0.35-0.57) | 3.55E-10 |
| 10-20 | 0.67 (0.57-0.79) | 1.78E-06 | 0.51 (0.43-0.60) | 1.37E-14 | 0.63 (0.53-0.74) | 2.36E-08 | 0.59 (0.49-0.70) | 4.37E-09 |
| 20-40 | 0.86 (0.75-0.98) | 0.02 | 0.63 (0.55-0.72) | 2.05E-11 | 0.72 (0.63-0.82) | 4.53E-07 | 0.81 (0.70-0.92) | 1.40E-03 |
| 40-60 | 1.00 (Reference) | - | 1.00 (Reference) | - | 1.00 (Reference) | - | 1.00 (Reference) | - |
| 60-80 | 1.45 (1.27-1.64) | 1.61E-08 | 1.37 (1.21-1.56) | 9.54E-07 | 1.03 (0.91-1.16) | 0.65 | 1.34 (1.18-1.51) | 5.33E-06 |
| 80-90 | 1.87 (1.60-2.19) | 6.92E-15 | 1.77 (1.52-2.07) | 1.88E-13 | 1.39 (1.21-1.61) | 6.30E-06 | 1.70 (1.47-1.97) | 1.15E-12 |
| 90-95 | 2.30 (1.88-2.80) | 1.94E-16 | 2.48 (2.06-3.00) | 2.46E-21 | 1.54 (1.29-1.85) | 2.47E-06 | 2.08 (1.74-2.49) | 1.44E-15 |
| >95 | 3.39 (2.80-4.10) | 1.16E-35 | 5.22 (4.37-6.24) | 7.13E-74 | 2.23(1.87-2.65) | 1.14E-19 | 3.98 (3.37-4.71) | 3.31E-58 |

PRS, polygenic risk score; IRS, integrated risk score; OR, odds ratio; CI, confidence interval; AUC, aera under the receiver operating characteristic curve.

[a] Among the whole ABCC datasets, except for those using Exome BeadChip or iCOGs as genotyping platform, 10,207 Chinese women who hx`ad both individual genetic and nongenetic data available were eligible for this analysis.

[b] $PRS_{111}$: the best PRS derived in the present study. $IRS_{111}$: the combination of $PRS_{111}$ and the NgRS.

[c] $PRS_{263\text{-}META}$ was derived based on the 330-SNP European PRS reported by Zhang et al. *Nat Genet*. 2020. $IRS_{263\text{-}META}$ was the combination of $PRS_{263\text{-}META}$ and the NgRS.

[d] OR and 95% CI of each PRS/IRS percentile group compared with the reference group and P values were estimated using logistic regression.
